# Integration of biomarker polygenic risk score improves prediction of coronary heart disease in UK Biobank and FinnGen

**DOI:** 10.1101/2022.08.22.22279057

**Authors:** Jake Lin, Nina Mars, Yu Fu, Pietari Ripatti, Tuomo Kiiskinen, FinnGen, Taru Tukiainen, Samuli Ripatti, Matti Pirinen

**Affiliations:** Institute for Molecular Medicine Finland FIMM, Helsinki Institute of Life Science HiLIFE, University of Helsinki, Helsinki, Finland; Department of Public Health, University of Helsinki, Helsinki, Finland; Massachusetts General Hospital & Broad Institute of Massachusetts Institute of Technology and Harvard University, Cambridge, MA, USA; Department of Mathematics and Statistics, University of Helsinki, Helsinki, Finland

## Abstract

**Background:** In addition to age and sex, also smoking history and levels of blood pressure, cholesterol, lipoproteins and inflammation are established biomarkers for coronary heart disease (CHD). As standard polygenic risk scores (PRS) have recently proven successful for CHD prediction, it remains of high interest to determine how a combined PRS of biomarkers (BioPRS) constructed from statistically relevant biomarkers can further improve genetic prediction of CHD.

**Methods:** We developed CHDBioPRS, which combines BioPRS with PRS of CHD, via regularized regression in UK Biobank (UKB) training data (*n* = 208,010). The resulting CHDBioPRS was tested on an independent UK Biobank subset (*n* = 25,765) and on the FinnGen study (*n* = 306,287).

**Results:** We observed a consistent pattern across all data sets where BioPRS was clearly predictive of CHD and improved standard PRS for CHD when the two were combined. In UKB test data, CHDPRS had a hazard ratio (HR) of 1.78 (95% confidence interval 1.67-1.91, area under the curve (AUC) 0.808) and CHDBioPRS had a HR of 1.88 (1.75-2.01, AUC 0.811) per one standard deviation of PRS. In FinnGen data, HR of CHDPRS was 1.57 (1.55-1.60, AUC 0.752) and HR of CHDBioPRS was 1.60 (1.58-1.62, AUC 0.755). We observed larger effects of CHDBioPRS in subsets of early onset cases with HR of 2.07 (1.85-2.32, AUC 0.790) in UKB test data and of 2.10 (2.04-2.16, AUC 0.791) in FinnGen. Results were similar when stratified by sex.

**Conclusions:** Integration of biomarker based BioPRS improved on the standard PRS for CHD and the gain was largest with early onset CHD cases. These findings highlight the benefit of enriching polygenic risk prediction of CHD with the genetics of associated biomarkers.

## BACKGROUND

Coronary heart disease (CHD), a complex disease caused by a gradual build-up of fatty deposits in the arteries, is a major cause of death worldwide. In addition to family history, age, sex, smoking history, and levels of blood pressure, inflammation and lipoproteins are established biomarkers for CHD (Visseren et al. 2021; Sinnott-Armstrong et al. 2021; Johnston et al. 2006; Zakynthinos and Pappa 2009). These risk factors are also used in clinical risk calculators to evaluate preventive therapies and strategies. While clinical risk scores enable identification of some individuals at high risk (Wilson, Castelli, and Kannel 1987; D’Agostino et al. 2008; Mahmood et al. 2014), a large proportion of CHD cases are not detected by these scores and the utility of clinical scores is limited for young adults (Wachira and Stys 2013; Johnston et al. 2006; McMahan et al. 2005; Aggarwal, Srivastav; Berry et al. 2007) and for women (Aggarwal et al. 2015; Garcia et al. 2016).

Genome-wide association studies (GWAS) involving large human genetic data sets have identified more than 100 loci statistically associated with CHD, mostly within populations of European descent (Nikpay et al. 2015; van der Harst and Verweij 2018; Matsunaga et al. 2020; Schunkert et al. 2011; CARDIoGRAMplusC4D Consortium et al. 2013; Nelson et al. 2017). These genetic discoveries together with the introduction of sophisticated statistical tools that incorporate linkage disequilibrium information, have greatly advanced risk prediction (Vilhjálmsson et al. 2015; Privé, Arbel, and Vilhjálmsson 2020; Ge et al. 2019). Particularly, several studies have shown that inclusion of polygenic risk scores (PRSes) improve CHD prediction and identification of high-risk groups (Abraham et al. 2016; Nikpay et al. 2015; Khera et al. 2018; Ripatti et al. 2010). Because PRSes are based on germline DNA, risk profiling can be conducted in early life when the individuals with the highest values of PRS are likely to benefit from an early adoption of preventive strategies.

The landmark study of PRSes for common diseases conducted by Khera et al. (Khera et al. 2018) showed that a sizable portion of the population carry a polygenic CHD risk equivalent to known monogenic mutations conferring several-fold increased risk. This PRS, comprising more than 6 million SNPs, was generated using LDPred (Vilhjálmsson et al. 2015) and has proven effective in validation sets across multiple populations while also performing favorably compared to other polygenic risk scores (Wünnemann et al. 2019) composed of smaller numbers of variants.

As PRSes have proven successful for CHD prediction, it remains of high interest to systematically determine how a combined biomarker score (BioPRS) constructed from biomarkers associated with CHD can improve upon the established CHDPRS. Recently, multi-PRS models, using 35 PRSes from blood and urine biomarkers, have been shown to improve genetic risk prediction of common disease such as type 2 diabetes and gout (Sinnott-Armstrong et al. 2021). While other existing studies have focused on combining several GWAS of CHD (Inouye et al. 2018; Munz et al. 2018; Nikpay et al. 2015), our focus is on the combination of effects of known CHD associated biomarkers into a single PRS and its integration with CHDPRS. Furthermore, as the current risk calculators do not work equally well for women as for men, it is of high importance to quantify the contribution of BioPRS within each sex (Brewer, Svatikova, and Mulvagh 2015). Another important goal is to predict a subgroup of CHD cases with an early onset of the disease (Mars et al. 2020).

## METHODS

The workflow of the study is in Figure 1.

**Figure 1.**
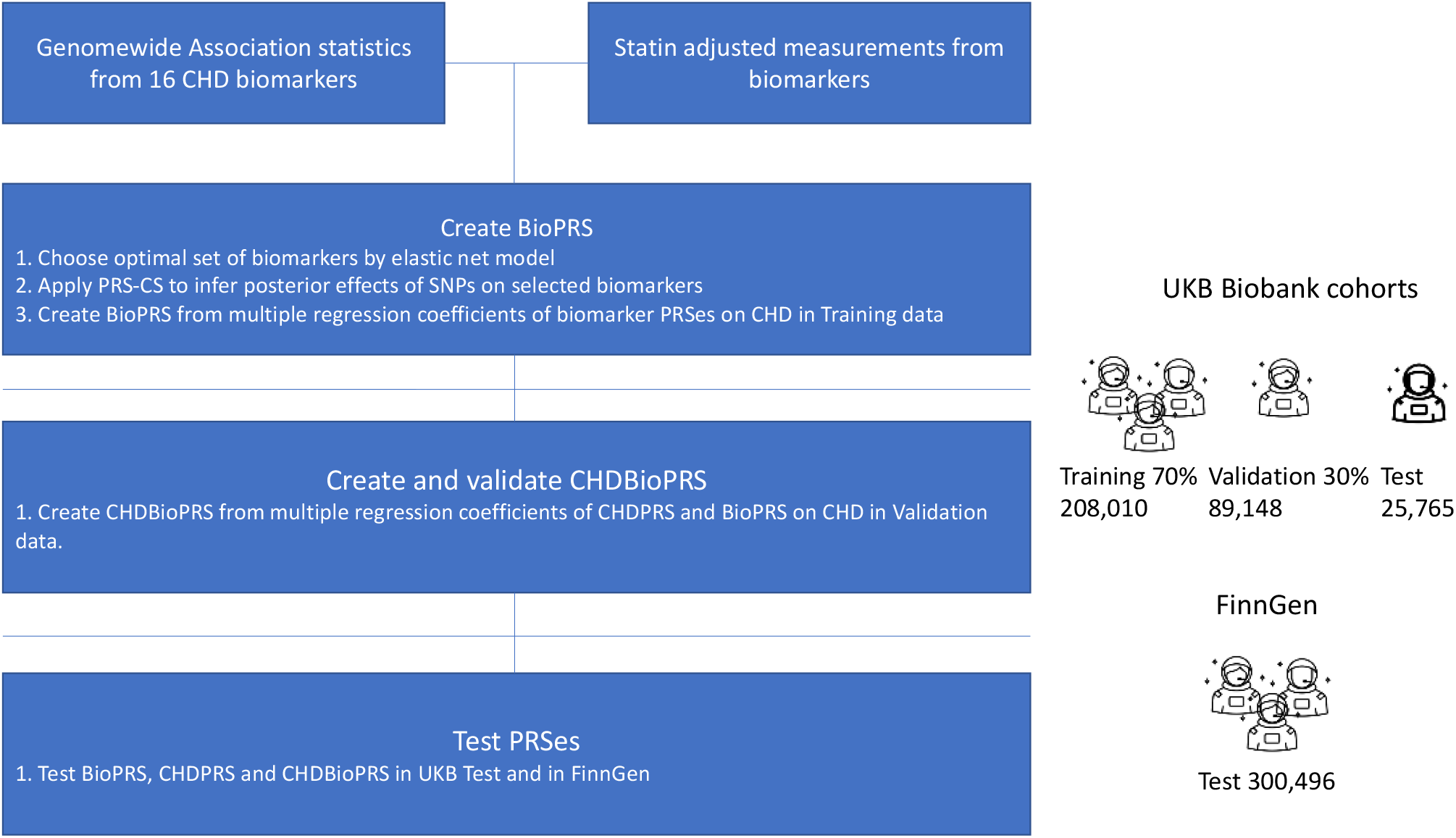
Study design and workflow. **CHD biomarkers**: We identified CHD associated risk factors or biomarkers from UKB (Table 2 and Supplementary Table S1). Genome-wide association study (GWAS) was performed on UKB genotype data, separately in females and males, for 16 biomarkers. **Construction of CHDBioPRS**: We regressed CHD on the biomarkers in the UKB training data using elastic net Cox regression and retained 10 biomarkers. BioPRS is constructed by weighting each PRS by its coefficient in joint Cox regression model for CHD in UKB training data. CHDBioPRS is the sum of the standard CHDPRS and BioPRS where the weights are estimated from a Cox regression model predicting CHD within UKB validation data.

**Figure 2.**
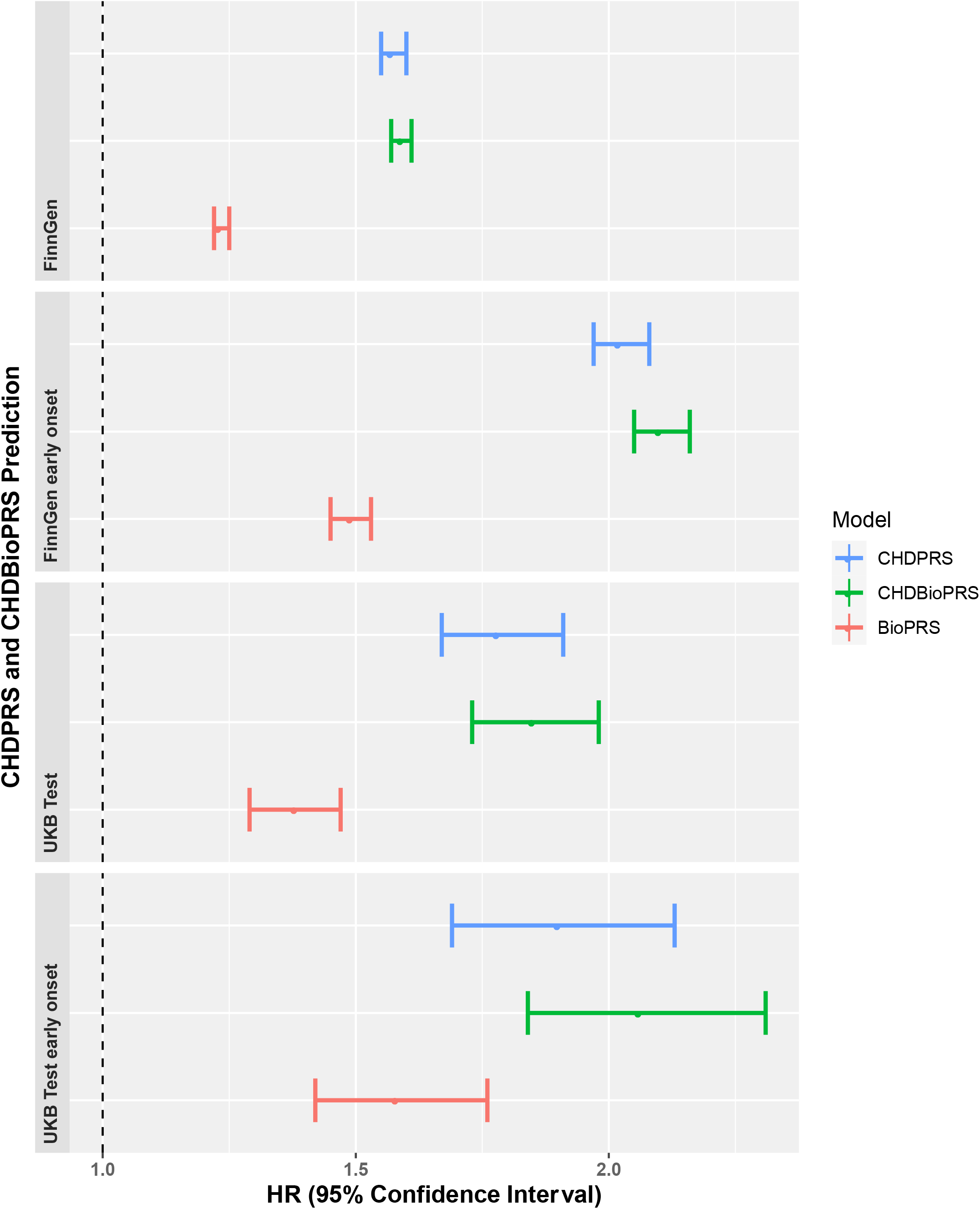
CHD prediction using PRSes in FinnGen and UKB Test samples. Both cohorts are disjoint from the samples used in construction of BioPRS and CHDBioPRS. Early onset = onset < 55 years of age.

### UKB data

The design of UKB and the background of its participants have been reported previously (Sudlow et al. 2015; Abraham et al. 2016). We restricted our analyses to samples that were of self-reported European ancestry to avoid potential spurious associations driven by allele frequency differences when including individuals from different ancestry backgrounds in our GWAS. We made use of the following four sets of UKB samples. First, our GWAS set contained 343,695 samples who were unrelated (pairwise kinship coefficients reported by UKB < 0.044) and who self-reported “white British” as their ethnicity. Out of the GWAS set, 297,158 individuals had data on all 16 biomarkers (listed below) and we split these into our UKB training (70%, 208,010 individuals) and UKB validation (30%, 89,148 individuals) sets. The split was done randomly by maintaining a constant CHD case ratio and sex ratio within each set. Finally, we also collected a UKB test set of 25,765 unrelated individuals with self-reported ethnicity as “white non-British” and biomarker measurements available. The UKB test set did not overlap with the UKB GWAS set, UKB training set or UKB validation set.

### UKB CHD definition

The UKB CHD endpoint was defined as fatal or nonfatal myocardial infarction (MI), or coronary revascularization (percutaneous transluminal coronary angioplasty, PTCA, or coronary artery bypass grafting, CABG). In detail, and consistent with previous studies (Khera et al. 2018; Tamlander et al. 2022), CHD cases were defined as having a heart attack diagnosed by a doctor (field 6150) or self-reported as a non-cancer illness (field 20002) or operations including PTCA and CABG (field 20004). In addition, coronary revascularization was assigned based on OPCS-4 (field 41272) coded procedure for CABG (K40.1-4, K41.1-4, and K45.1-5) or PTCA (K49.1-2, K49.8-9, K50.2, K75.1-4 and K75.8-9). MI cases from hospital episode statistics (fields 41202 (ICD-10) and 41203 (ICD-9)) were defined with ICD-10 I21, I22, I23, I24.1 and I25.2 and ICD-9 codes of 410, 411 and 412. We further merged the outcomes from the UKB MI algorithmically-defined outcomes and dates (fields 42000 and 42001). We defined prevalent CHD cases as those who had CHD already when their blood sample was taken; other CHD cases were considered incident CHD cases. The age at onset in prevalent cases was determined by hospital episode data and self-reported age of onset while for incident CHD cases by hospital or death records demonstrating disease onset after UKB enrollment.

### UKB Biomarkers and GWAS

We identified 16 variables from UKB that have been previously reported associated with CHD (Table 2 and Supplementary Table S1): HDL cholesterol (field 30760), LDL direct (30780), Triglycerides (TRIG, 30870), ApoA1 (30630), ApoB (30640), diastolic blood pressure (DBP, 4079), systolic blood pressure (SBP, 4080), glycated hemoglobin (HbA1c, 30750), glucose (30740), C-reactive protein (CRP, 30710), creatinine (CREA, 30700), lipoprotein(a) (LPA, 30790) doctor diagnosed diabetes (2443), body mass index (BMI, 21001) and cigarettes per day (CPD, 3456). In addition, total cholesterol (TC) was calculated from the Friedewald formula (Friedewald, Levy, and Fredrickson 1972) as HDL + LDL + TG/2.2 in units of mmol/L. Following the previous biomarker study by Sinnot-Armstrong et al. (Sinnott-Armstrong et al. 2021), statin usage is identified from treatment/medication (field 20003, 13 drugs: 1141146234, atorvastatin; 1141192414, crestor 10mg tablet; 1140910632, eptastatin; 1140888594, fluvastatin; 1140864592, lescol 20 mg capsule; 1141146138, lipitor 10mg tablet; 1140861970, lipostat 10mg tablet; 1140888648, pravastatin; 1141192410, rosuvastatin; 1141188146, simvador 10mg tablet; 1140861958, simvastatin; 1140881748, zocor 10mg tablet; 1141200040, zocor heart-pro 10mg tablet). Adjustment for statin usage (Sinnott-Armstrong et al. 2021) was done for the following biomarkers by dividing the biomarker value with the given coefficient: HDL (1.053), LDL (0.684), TRIG (0.874), ApoA1 (1.07), ApoB (0.722), LPA (1.102), GLUC (1.029), CRP (1.230), CREA (1.058) and HbA1c (1.042). We also adjusted the subjects taking blood pressure reduction drugs (fields 6153 and 6177) by adding +15 to SBP and +10 to DBP. The statin adjusted values of these biomarkers for the UKB training set are shown in Table 2.

**Table 1:**
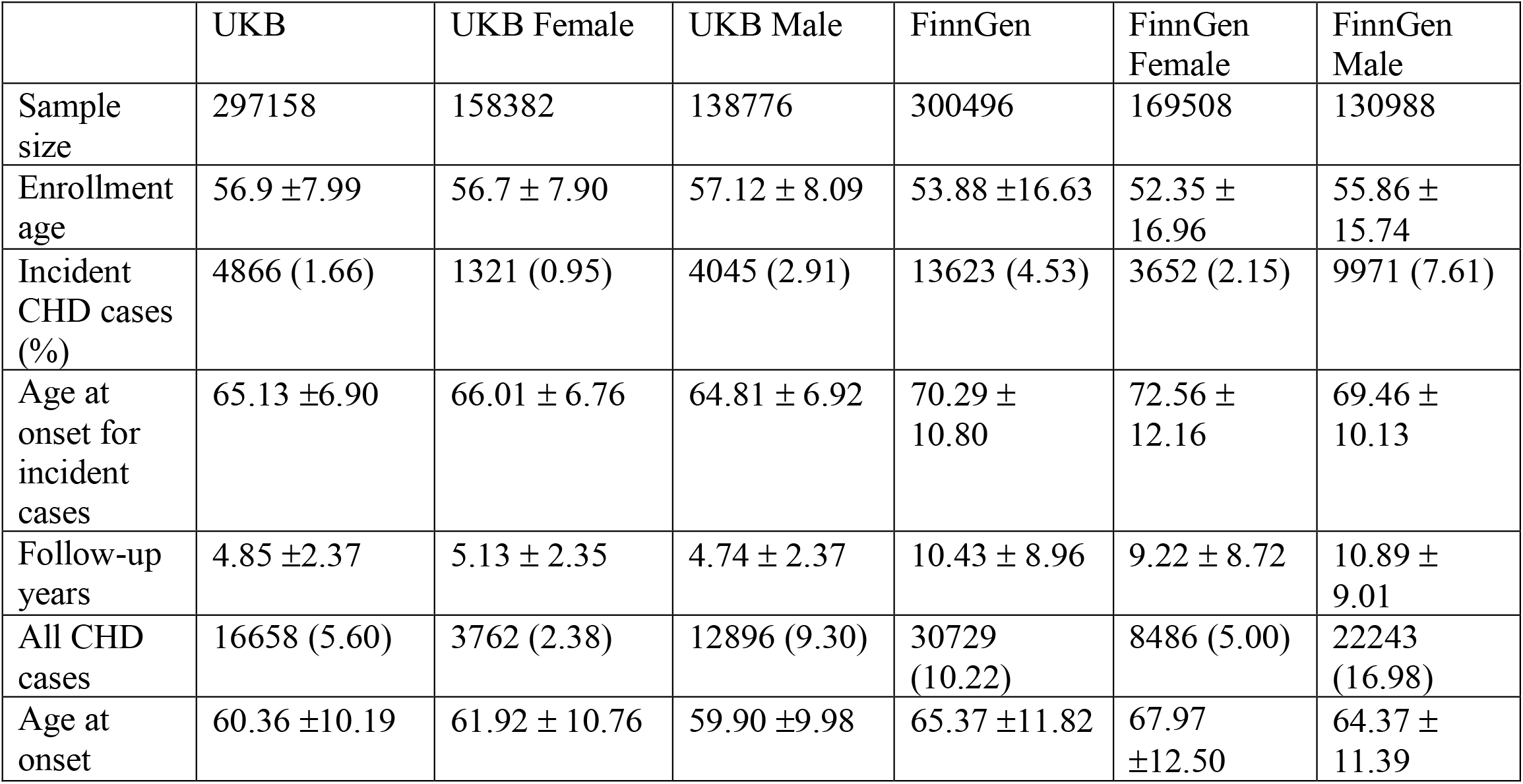
Sample characteristics in UK Biobank (UKB) and FinnGen. Values are either mean ± standard deviation or count (percentage). For incident CHD cases, age at onset is after the blood sample was taken.

**Table 2:**
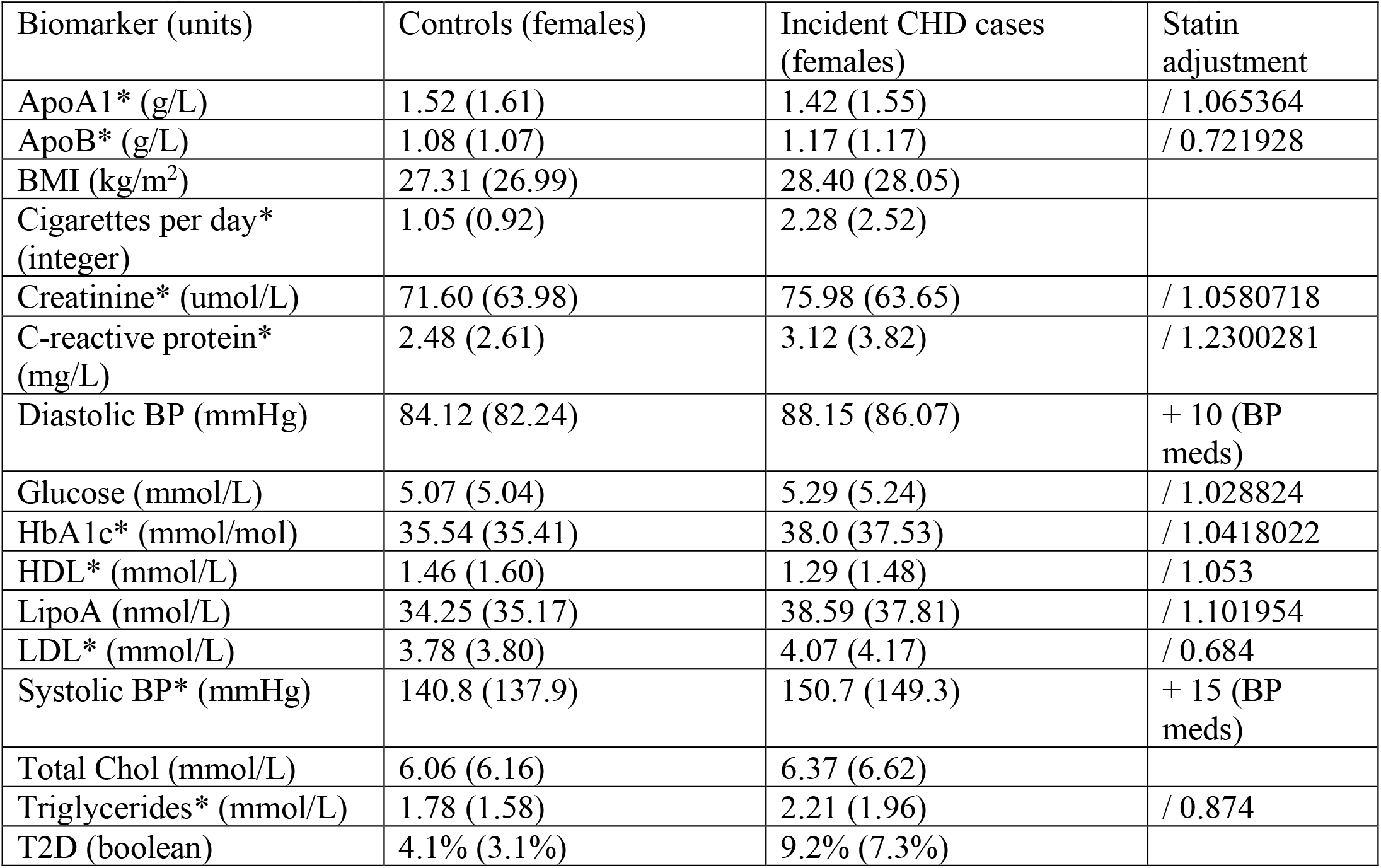
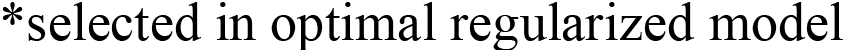
UKB Biomarkers mean levels after adjustment for statins. Statin adjustment is done either by division (/) or addition (+) by the value given in the last column for statin users (16.2%).

| Biomarker (units) | Controls (females) | Incident CHD cases (females) | Statin adjustment |
| --- | --- | --- | --- |
| ApoA1* (g/L) | 1.52 (1.61) | 1.42 (1.55) | / 1.065364 |
| ApoB* (g/L) | 1.08 (1.07) | 1.17 (1.17) | / 0.721928 |
| BMI (kg/m<sup>2</sup>) | 27.31 (26.99) | 28.40 (28.05) |  |
| Cigarettes per day* (integer) | 1.05 (0.92) | 2.28 (2.52) |  |
| Creatinine* (umol/L) | 71.60 (63.98) | 75.98 (63.65) | / 1.0580718 |
| C-reactive protein* (mg/L) | 2.48 (2.61) | 3.12 (3.82) | / 1.2300281 |
| Diastolic BP (mmHg) | 84.12 (82.24) | 88.15 (86.07) | + 10 (BP meds) |
| Glucose (mmol/L) | 5.07 (5.04) | 5.29 (5.24) | / 1.028824 |
| HbA1c* (mmol/mol) | 35.54 (35.41) | 38.0 (37.53) | / 1.0418022 |
| HDL* (mmol/L) | 1.46 (1.60) | 1.29 (1.48) | / 1.053 |
| LipoA (nmol/L) | 34.25 (35.17) | 38.59 (37.81) | / 1.101954 |
| LDL* (mmol/L) | 3.78 (3.80) | 4.07 (4.17) | / 0.684 |
| Systolic BP* (mmHg) | 140.8 (137.9) | 150.7 (149.3) | + 15 (BP meds) |
| Total Chol (mmol/L) | 6.06 (6.16) | 6.37 (6.62) |  |
| Triglycerides* (mmol/L) | 1.78 (1.58) | 2.21 (1.96) | / 0.874 |
| T2D (boolean) | 4.1% (3.1%) | 9.2% (7.3%) |  |

\*selected in optimal regularized model

**Genome-wide association study (GWAS)**, using BOLT-LMM v2.3.2 (Loh et al. 2015) was performed on UKB genotype data (release 3), separately in 183,130 women and 157,821 men, for the 16 biomarkers. We first regressed out sex, age, age^2^ and the top 10 principal components of genetic structure from the biomarkers and then applied rank-based inverse-normal transformation to the residuals.

### FinnGen

The design of the Finnish FinnGen ((FinnGen 2021)) project and participant backgrounds are shown in Table 1. The FinnGen test cohort contained 321,302 FinnGen data freeze 7 (DF7) participants. The CHD case definition in FinnGen (I9_CHD) is consistent with our UKB CHD definition except that the FinnGen definition also includes samples with angina only (I20.0) as cases. Subsequently, we removed the 2,989 angina only cases from FinnGen, in addition to removal of 6,109 participants less than 16 years of age at enrollment.

### Biomarker model

Using as predictors the 16 CHD associated biomarkers for the UKB training data and incident CHD as outcome (and excluding prevalent CHD cases), we applied elastic net Cox regression as implemented in glmnet (Friedman et al. 2021; Hastie and Qian 2014) with alpha = 0.50 and using 20-fold cross validation. We chose the optimal model by glmnet’s “lambda.1se” criterion and identified 10 biomarkers (Table 2). As shown in the Supplementary Figure 1, an alpha = 0.25 produced the same optimal set as 0.50 setting while alpha = 0.75 further excluded ApoA1. The identified 10-biomarker model from lambda.1se was selected for BioPRS construction.

### Biomarker PRSes

PRS-CS (Ge et al. 2019) was run on each of the GWAS summary results of the selected 10 biomarkers. To account for linkage disequilibrium (LD), we employed the 1000 Genomes project’s (Nikpay et al. 2015; 1000 Genomes Project Consortium et al. 2015) phase 3 European reference panel which resulted in LD adjusted posterior effect sizes for 1,139,910 SNPs. The PRS for each biomarker was then computed as a sum over SNPs of the products of the individual’s genotype and the posterior effect size for the SNP using PLINK2.0 (Purcell and Chang, 2019).

### Weights of biomarker PRSes on CHD

The PRSes of the 10 selected biomarkers were used as predictors for incident CHD in a Cox proportional hazards model in the UKB training data without the prevalent CHD cases. According to this model, the hazard rate at age t depends on the predictors as follows:

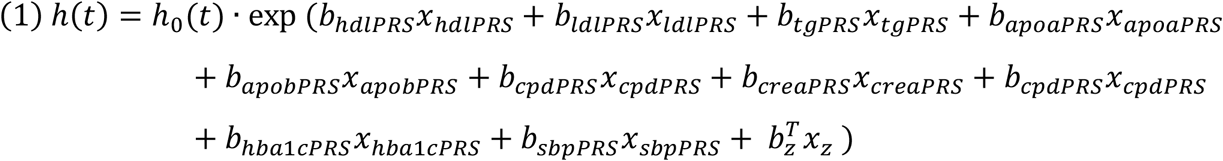

Here *h*_*0*_*(t)* is the baseline hazard rate, **z** denotes the vector of covariate values (sex, and the first 10 principal components of population structure) and each biomarker has coefficient *b*_biomarkerPRS_ that corresponds to a change in the logarithm of the hazard rate per one standard deviation of the biomarker PRS value.

We used the ‘coef’ function from the ‘survival’ library (Therneau 2021) of R software to estimate the *b*_biomarkerPRS_ coefficients as previously recommended (Fieuws and Verbeke 2004; Verbeke 1997).

### BioPRS

We combined the biomarker PRSes into a score named BioPRS by standardizing (mean of 0 and SD of 1) the sum of the 10 biomarker PRSes after multiplying each PRS by the beta coefficient (b_i_) of the corresponding biomarker from formula 1:

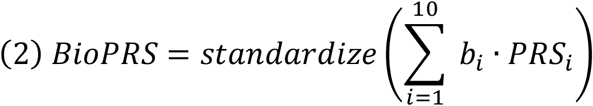

### CHDPRS

We generated a PRS for CHD (named CHDPRS) by applying PRS-CS (Ge et al. 2019) to CHD GWAS reported by Nikpay et al. (2015) using European panel from the 1000 Genomes Project (1000 Genomes Project Consortium et al. 2015) for LD reference. Our CHDPRS contained 1,087,715 SNPs. Additionally, we compared our CHDPRS to “Khera PRS” that is the PRS for CHD generated using LDpred by Khera et al. (2018) based on the same GWAS (Nikpay et al. 2015) that we used to generate our CHDPRS.

**CHDBioPRS** was constructed from integration of BioPRS and CHDPRS. Weights of the two PRSes (CHDPRS and mBioPRS) were estimated in the UKB validation set using a Cox regression model with CHD as outcome:

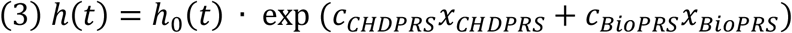

CHDBioPRS is the standardized sum of the CHDPRS and BioPRS multiplied by their weights from formula 3:

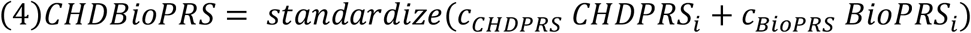

In addition to the derivation above, a similar procedure was done also for men and women separately (biomarker selection using glmnet, biomarker weights in BioPRS using Cox regression and combination of CHDPRS and BioPRS using another Cox regression).

### Early onset

We identified all individuals with early CHD onset (< 55 years) and, to further account for sex differences, we defined early CHD onset for women as < 60 years of age and early CHD onset for men as < 50 years of age.

### Statistical analysis

All scores were standardized to have a mean of 0 and a variance of 1. Hazard ratio (HR) and its 95% confidence interval were computed using ‘coxph’ from the survival package. In Cox regression, age was used as the time scale. When evaluating PRSes, the CHD outcome variable included both incident and prevalent cases. In UKB, we excluded prevalent cases from the analysis when we selected relevant biomarkers and when we estimated the biomarker weights for generating BioPRS. In survival analyses of early onset cases, the cases with late onset were excluded and the controls were censored at the upper limit of the early onset age.

C-Index (Harrell et al. 1982), a metric for prediction concordance, is obtained from the ‘survival’ (Therneau 2021) package in R software and Area under the curve (AUC) were used to assess model discrimination. AUC and its confidence interval were obtained from logistic regression (‘glm’ function) and ‘pROC’ package (Robin et al. 2011) in R software. We also computed how much each biomarker PRS explains of the variance of the corresponding biomarker by using the adjusted R^2^ measure as reported by ‘lm’ function in R software, for the linear model where the biomarker PRS was the only predictor in the model. This analysis was done in the UKB test data.

## RESULTS

For our UKB GWAS set, we generated statin adjusted GWAS results for 16 CHD associated biomarkers. After accounting for withdrawals and missing biomarker measurements, we identified a total of 297,158 participants (53.3% female) comprising of 16,658 CHD cases (22.6% female). We further split this data into disjoint training (70%) and validation (30%) sets while maintaining similar case and sex proportions. Our UKB test data set of 25,765 (56.4% female) unrelated individuals of “non-British white” self-identified ethnicity included 949 CHD cases (23.2% female). Our FinnGen test data contained 300,496 (56.4% women; 10.4% CHD cases of whom 27.6% women) Finnish individuals. A combined PRS of CHD biomarkers (BioPRS) was derived across PRSes of 10 biomarkers (HDL, LDL, TRIG, Total cholesterol, systolic blood pressure, cigarettes per day, HbA1c, C-Reactive protein, creatinine, ApoB, ApoA1) by combining the individual biomarker PRSes generated with PRS-CS on 1,106,191 SNPs. This BioPRS was further integrated with the CHDPRS constructed from 6,630,150 variants from a GWAS (Nikpay et al. 2015) involving 184,305 participants of European ancestry to yield CHDBioPRS. We also compared our CHDPRS to the Khera PRS (Khera et al. 2018) and saw that our CHDPRS gave slightly more predictive result in UKB training data (HR of 1.64, 95%CI (1.61-1.67) compared to 1.62 (1.59-1.64) (Supplementary Table S2)). CHDBioPRS is approximately normally distributed and, on average, higher in CHD cases relative to the controls (mean difference 0.518 SD, with 95% CI (0.490,0.546); Supplementary Figure S2).

Table 3 shows the results from UKB data sets and FinnGen. In UKB, results from the training set are similar to results from validation and test sets, which suggests that the model is not overfitting in the training data. In all three UKB data sets, we observe the same pattern where BioPRS itself is clearly predictive of CHD (HR estimates per standard deviation vary between 1.42-1.45), CHDPRS on its own is more predictive than BioPRS (HR estimates vary between 1.62-1.78) and CHDBioPRS is the most predictive (HRs vary between 1.73-1.88). For C-Index, Z-scores and AUC metrics see Supplementary Tables S2, S3 and S4. When the scores are applied in FinnGen (Supplementary Tables S5), we repeat the pattern where BioPRS is clearly predictive and improves the prediction by CHDPRS when combined with CHDPRS into CHDBioPRS. Overall, the HRs in FinnGen are smaller than in UKB for all PRSes.

**Table 3:** Hazard ratios (with 95% confidence intervals) from Cox regression model of three different polygenic risk scores (PRS) on incident CHD. BioPRS is combination of PRSes of 10 CHD related biomarkers, CHDPRS is a standard PRS for CHD, CHDBioPRS combines BioPRS and CHDPRS. Early onset was defined as CHD below 55 years of age. C-index, AUC and P-values are in Supplementary Tables S2-S7.

| **Cohort** | **BioPRS** | **CHDPRS** | **CHDBioPRS** |
| --- | --- | --- | --- |
| UKB Training | 1.45 (1.42-1.48) | 1.64 (1.61-1.67) | 1.76 (1.72-1.79) |
| UKB Validation | 1.42 (1.38-1.46) | 1.62 (1.57-1.67) | 1.73 (1.68-1.78) |
| UKB Test | 1.45 (1.36-1.55) | 1.78 (1.67-1.91) | 1.88 (1.76-2.01) |
| UKB Test females | 1.53 (1.34-1.75) | 1.72 (1.50-1.98) | 1.86 (1.62-2.14) |
| UKB Test males | 1.42 (1.32-1.53) | 1.72 (1.60-1.85) | 1.84 (1.71-1.98) |
| UKB Test early onset | 1.60 (1.43-1.78) | 1.90 (1.69-2.13) | 2.07 (1.85-2.32) |
| FinnGen | 1.27 (1.26-1.29) | 1.57 (1.55-1.60) | 1.60 (1.58-1.62) |
| FinnGen females | 1.27 (1.24-1.30) | 1.53 (1.50-1.57) | 1.56 (1.53-1.60) |
| FinnGen males | 1.27 (1.25-1.29) | 1.58 (1.55-1.60) | 1.61 (1.59-1.63) |
| FinnGen early onset | 1.51 (1.47-1.55) | 2.01 (1.95-2.07) | 2.10 (2.04-2.16) |

By comparing the CHD risk between the top 5% of the PRS distribution and the rest, we observed larger HRs for CHDBioPRS (UKB Test 4.16 (3.09-5.60), FinnGen 4.04 (3.76-4.35)) than for CHDPRS (UKB Test 3.56 (2.62-4.86), FinnGen 3.84 (3.56-4.14), HRs for other percentiles along with their AUCs are listed in Supplementary Tables S6 for UKB Test and S7 for FinnGen).

We next studied the PRSes in cases with early CHD onset (< 55 years). For UKB test data, we had 325 cases (females 20.6%) and for FinnGen, 5965 cases (21.4% female). For both cohorts, we observed higher HRs for early onset cases than for all cases and again we saw HRs growing when using CHDBioPRS (UKB test 2.07 (1.85-2.32), FinnGen 2.10 (2.04-2.16)) instead of CHDPRS (UKB test 1.90 (1.69-2.13, FinnGen 2.01 (1.95-2.07) Table 3; see also Supplementary Tables S6 for UKB Test and S7 for FinnGen). Among all CHD cases, the CHDBioPRS values peaked for cases with an onset at around 40 years of age (Supplementary Figure S3).

It is known that CHD incidence and biomarker associations vary between sexes (Johnston et al. 2006). Therefore, we stratified UKB data and FinnGen data by sex. In both sexes, the regularized optimal models built in UKB training data consisted of the same 8 biomarkers (HDL, LDL, TRIG, SBP, CPD, HbA1c, CRP, ApoB). Using these biomarkers, we refitted the models in each sex separately and derived corresponding scores (BioPRS and CHDBioPRS). The CHD HR had similar dynamics between CHDPRS and CHDBioPRS compared to the full cohort (Table 3 and Supplementary Tables S8-S11). Among FinnGen women with onset below 60 years of age, the HRs were 1.82 (1.74-1.90) for CHDPRS and 1.90 (1.81-1.99) for CHDBioPRS (Supplementary Table S12) and for FinnGen males with onset below 50 years of age, the HRs were 2.17 (2.07-2.27) for CHDPRS and 2.26 (2.16-2.36) for CHDBioPRS (Supplementary Table S13).

## DISCUSSION

In an analysis of more than 600,000 participants involving two nationwide study cohorts, UK biobank and FinnGen, we showed that combining a biomarker polygenic risk score derived from 10 CHD associated biomarkers with a standard PRS for CHD improved on prediction of CHD. This improvement in prediction was largest for the early-onset CHD for both men and women.

Our BioPRS compared well with the recently published multi-PRS score by Sinnott-Armstrong et al. (Sinnott-Armstrong et al. 2021) that was derived from 35 UKB biomarkers. Their multi-PRS score was reported to have a HR of 1.19 (1.17-1.22) in FinnGen (data freeze 3) for myocardial infarction (Sinnott-Armstrong et al. 2021) while our BioPRS achieved HR of 1.29 (1.27-1.31) using the same endpoint and same covariates in a more recent and larger data freeze 7 of FinnGen.

Additionally, when Sinnott-Armstrong et al. combined their multi-PRS with a standard PRS for CHD in FinnGen, they reported a HR of 1.50 (1.46-1.53) (Sinnott-Armstrong et al. 2021), while our CHDBioPRS achieved a HR of 1.60 (1.58-1.62) in FinnGen (data freeze 7). Similarly, a MetaGRS score from Inouye et al. (Inouye et al. 2018), that combined three genetic risk scores (GRS46K (Abraham et al. 2016), another score based on 202 significant genetic variants from CARDIOGRAMplusC4D (Nikpay et al. 2015) and a genome-wide score based on the same GWAS (Nikpay et al. 2015)), reported a HR of 1.71 (1.68-1.73) when tested on UKB while our CHDBioPRS yielded a HR of 1.88 (1.75-2.01) in our UKB Test data.

There were clear differences in effect sizes between UKB and FinnGen. These differences could in part relate to differences in sample ascertainment procedures and genetic background. The UKB participants are known to be healthier than the general population (Fry et al. 2017) and therefore the relative contribution of genetics to their disease risk may be larger, whereas the FinnGen participants have been recruited through their contacts with the Finnish healthcare system.

Additionally, the FinnGen participants are on average 5 years older compared to the UK biobank participants and the CHD case rate in FinnGen is nearly double compared to UK biobank (10.2% vs. 5.6%). The biomarker GWAS effect sizes and linkage disequilibrium information used in creating PRS were derived in UKB or from other non-Finnish European populations, which could lead to better predictive power of PRSes in UKB compared to Finnish data (Kerminen et al. 2019). For our CHDPRS, the effect sizes were taken from a large GWAS meta-analysis (Nikpay et al. 2015) that may have included some of our FinnGen test samples. However, since our CHDPRS performed better in UKB than in FinnGen, we do not expect this potential overlap to have caused serious overfitting in our FinnGen test data.

Both sex-specific CHDBioPRSes were constructed from the same set of 8 biomarkers (HDL, LDL, triglycerides, systolic blood pressure, cigarettes per day, HbA1c, CRP, ApoB) and achieved prediction improvements compared to the standard CHD PRS. Importantly, we observed fairly similar hazard ratio estimates for men and women, which is in contrast with existing CHD clinical scores, such as QRISK/QRISK2 that is known to underestimate the CHD risks in women (Saar et al. 2019; Sedak et al. 2020; Woodward 2019). We also observed larger hazard ratios for early onset cases than all cases indicating that our PRSes are also informative about age at onset.

Disease risk prediction using multiple biomarkers and genome-wide set of genetic variants is a very high-dimensional problem and therefore adding more sparsity to the model building could improve the risk prediction. For example, one could attempt to use, for each genomic region separately, only a relevant subset of biomarkers (Lin et al. 2020). Furthermore, genomic regions could be prioritized, for example, by curated CHD molecular pathways (Liberzon et al. 2015) including known lipid-associated genomic regions (Willer et al. 2013).

Our study is limited to individuals with European ancestry. Given recent discoveries about imperfect transferability of PRS between populations (Martin et al. 2019; Mars et al. 2022), training of an optimal CHDBioPRS for non-European ancestries would require appropriate training data from other ancestries. This is also important since the potential to use genetic scores to identify high risk individuals from birth could exacerbate the health differences between individuals with European ancestry and the others until a broader inclusion of underserved ethnicities in research become reality, particularly in multi-ethnic countries such as the UK and the US (Sirugo, Williams, and Tishkoff 2019).

An approach similar to our BioPRS could also improve prediction of other complex diseases, such as Type 2 diabetes and breast cancer, with established PRSes and known, heritable risk factors (Mars et al. 2020).

## CONCLUSION

The integration of biomarker PRSes improves on the standard PRS for prediction of CHD, where the gain was the largest among the early onset CHD cases. This study strengthens the evidence for genome-based CHD prediction and quantifies the interplay between standard CHD PRS and PRSes of biomarkers associated with CHD.

## Data Availability

All data produced in the present study are available upon reasonable request to the authors. Upon acceptance, all relevant data will be submitted to the Polygenic Score Catalog.

## Supplementary tables

https://docs.google.com/spreadsheets/d/1AR8lo_lrzcCJoXz_ETZuSLB5waXD6iM8/edit#gid=2034380250

## Ethics statement and materials & methods

Patients and control subjects in FinnGen provided informed consent for biobank research, based on the Finnish Biobank Act. Alternatively, separate research cohorts, collected prior the Finnish Biobank Act came into effect (in September 2013) and start of FinnGen (August 2017), were collected based on study-specific consents and later transferred to the Finnish biobanks after approval by Fimea (Finnish Medicines Agency), the National Supervisory Authority for Welfare and Health. Recruitment protocols followed the biobank protocols approved by Fimea. The Coordinating Ethics Committee of the Hospital District of Helsinki and Uusimaa (HUS) statement number for the FinnGen study is Nr HUS/990/2017.

The FinnGen study is approved by Finnish Institute for Health and welfare (permit numbers: THL/2031/6.02.00/2017, THL/1101/5.05.00/2017, THL/341/6.02.00/2018, THL/2222/6.02.00/2018, THL/283/6.02.00/2019, THL/1721/5.05.00/2019, THL/1524/5.05.00/2020, and THL/2364/14.02/2020), Digital and population data service agency (permit numbers: VRK43431/2017-3, VRK/6909/2018-3, VRK/4415/2019-3), the Social Insurance Institution (permit numbers: KELA 58/522/2017, KELA 131/522/2018, KELA 70/522/2019, KELA 98/522/2019, KELA 138/522/2019, KELA 2/522/2020, KELA 16/522/2020, Findata THL/2364/14.02/2020 and Statistics Finland (permit numbers: TK-53-1041-17 and TK/143/07.03.00/2020 (earlier TK-53-90-20).

The Biobank Access Decisions for FinnGen samples and data utilized in FinnGen Data Freeze 7 include: THL Biobank BB2017_55, BB2017_111, BB2018_19, BB_2018_34, BB_2018_67, BB2018_71, BB2019_7, BB2019_8, BB2019_26, BB2020_1, Finnish Red Cross Blood Service Biobank 7.12.2017, Helsinki Biobank HUS/359/2017, Auria Biobank AB17-5154 and amendment #1 (August 17 2020), Biobank Borealis of Northern Finland_2017_1013, Biobank of Eastern Finland 1186/2018 and amendment 22 § /2020, Finnish Clinical Biobank Tampere MH0004 and amendments (21.02.2020 & 06.10.2020), Central Finland Biobank 1-2017, and Terveystalo Biobank STB 2018001.

## Acknowledgements

This work was supported by the Academy of Finland (grant nos. 325999 for JL, 331671 for NM, 312076 and 336825 for MP, 285380 and 312062 for SR), by Sigrid Juselius Foundation (MP and SR).

We want to acknowledge the participants and investigators of FinnGen study. The FinnGen project is funded by two grants from Business Finland (HUS 4685/31/2016 and UH 4386/31/2016) and the following industry partners: AbbVie Inc., AstraZeneca UK Ltd, Biogen MA Inc., Bristol Myers Squibb (and Celgene Corporation & Celgene International II Sàrl), Genentech Inc., Merck Sharp & Dohme LCC, Pfizer Inc., GlaxoSmithKline Intellectual Property Development Ltd., Sanofi US Services Inc., Maze Therapeutics Inc., Janssen Biotech Inc, Novartis AG, and Boehringer Ingelheim International GmbH. Following biobanks are acknowledged for delivering biobank samples to FinnGen: Auria Biobank (www.auria.fi/biopankki), THL Biobank (www.thl.fi/biobank), Helsinki Biobank (www.helsinginbiopankki.fi), Biobank Borealis of Northern Finland (https://www.ppshp.fi/Tutkimus-ja-opetus/Biopankki/Pages/Biobank-Borealis-briefly-in-English.aspx), Finnish Clinical Biobank Tampere (www.tays.fi/en-US/Research_and_development/Finnish_Clinical_Biobank_Tampere), Biobank of Eastern Finland (www.ita-suomenbiopankki.fi/en), Central Finland Biobank (www.ksshp.fi/fi-FI/Potilaalle/Biopankki), Finnish Red Cross Blood Service Biobank (www.veripalvelu.fi/verenluovutus/biopankkitoiminta) and Terveystalo Biobank (www.terveystalo.com/fi/Yritystietoa/Terveystalo-Biopankki/Biopankki/). All Finnish Biobanks are members of BBMRI.fi infrastructure (www.bbmri.fi). Finnish Biobank Cooperative -FINBB (https://finbb.fi/) is the coordinator of BBMRI-ERIC operations in Finland. The Finnish biobank data can be accessed through the Fingenious® services (https://site.fingenious.fi/en/) managed by FINBB.

This research has been conducted using the UK Biobank Resource under Application Number 22627.

## Contributors of FinnGen

### Steering Committee

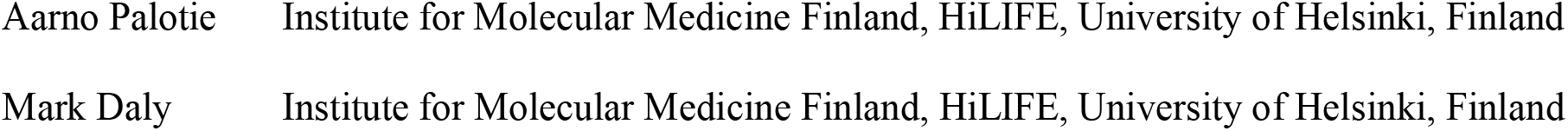

#### Pharmaceutical companies

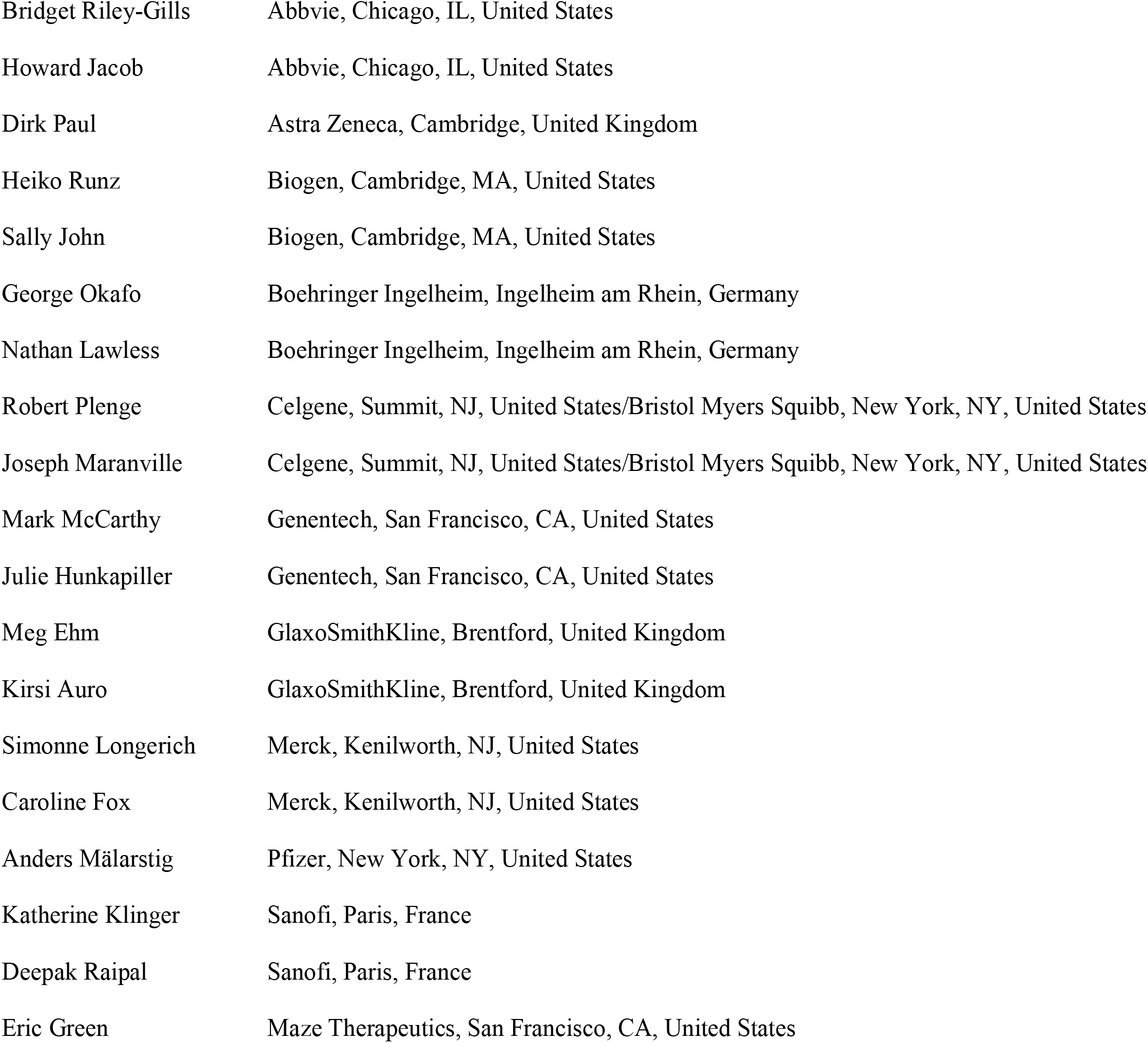
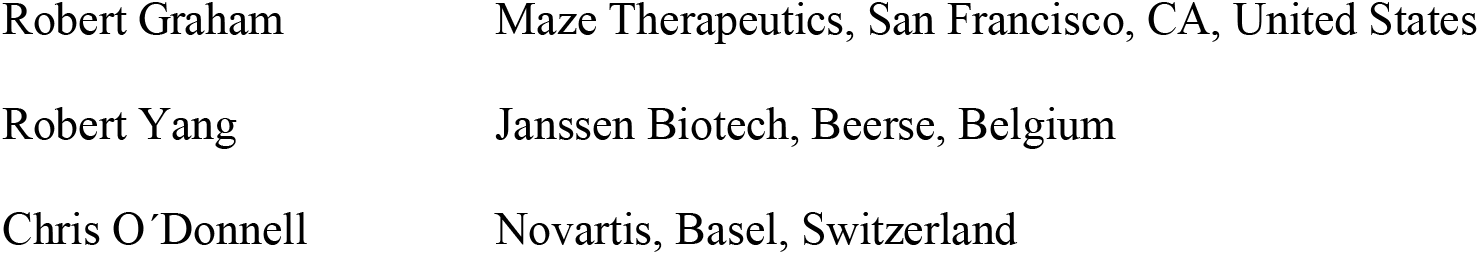

#### University of Helsinki & Biobanks

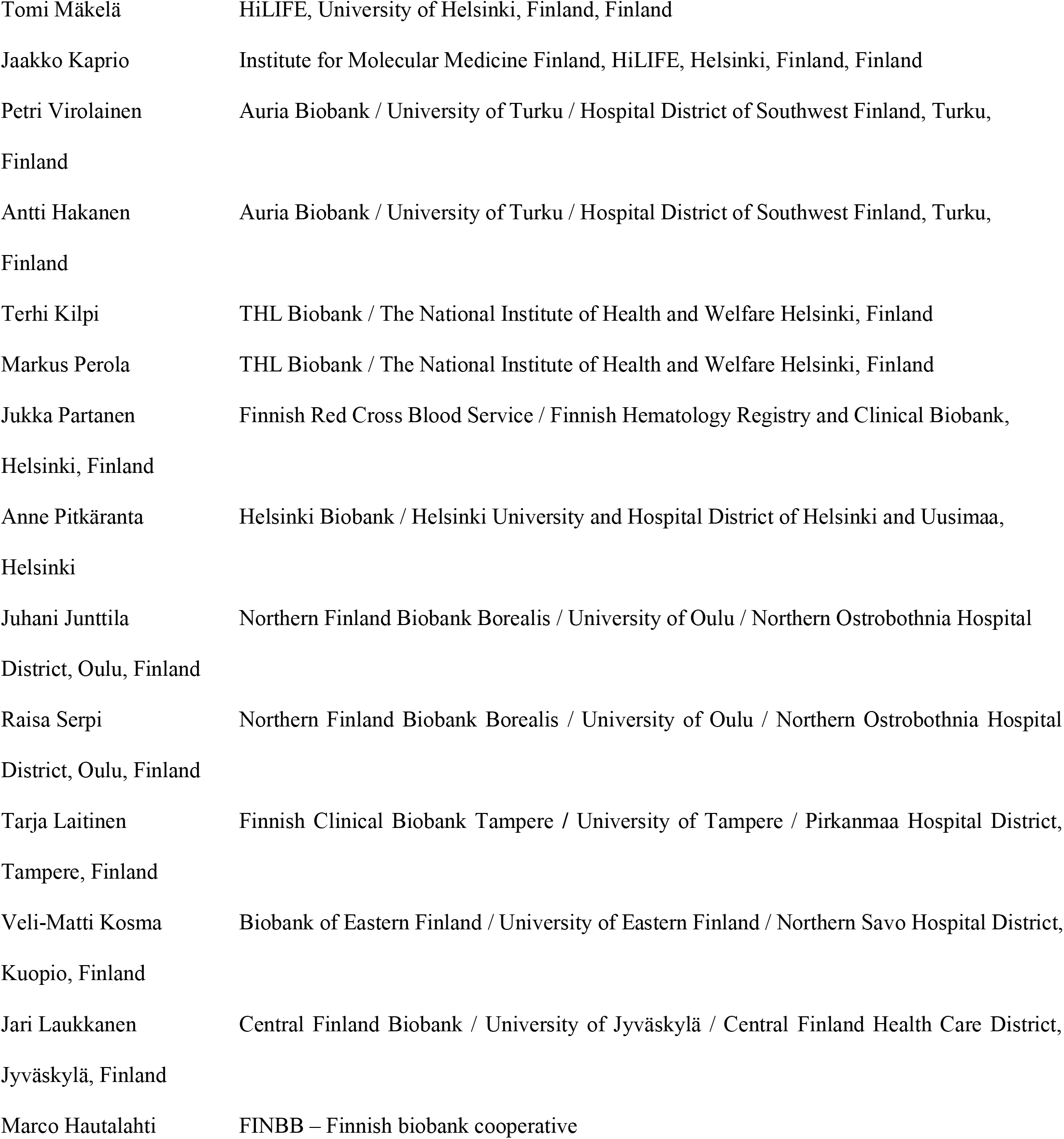

#### Other Experts/ Non-Voting Members

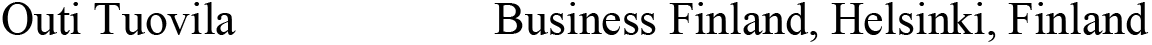
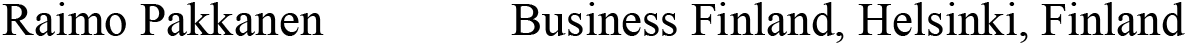

#### Scientific Committee

##### Pharmaceutical companies

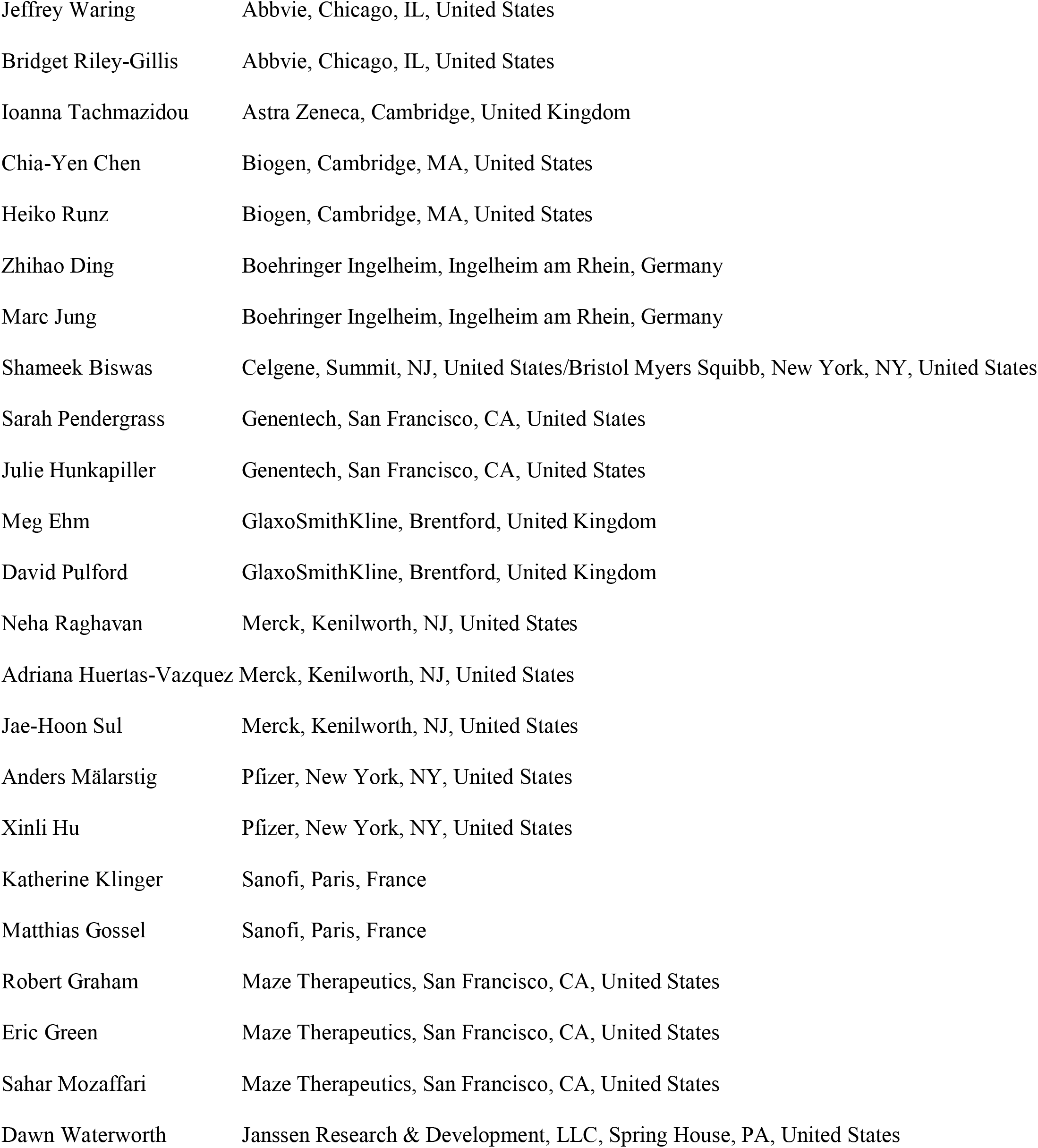
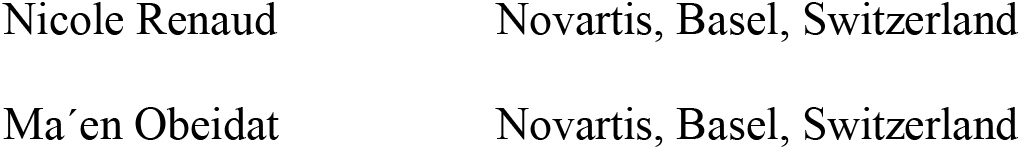

##### University of Helsinki & Biobanks

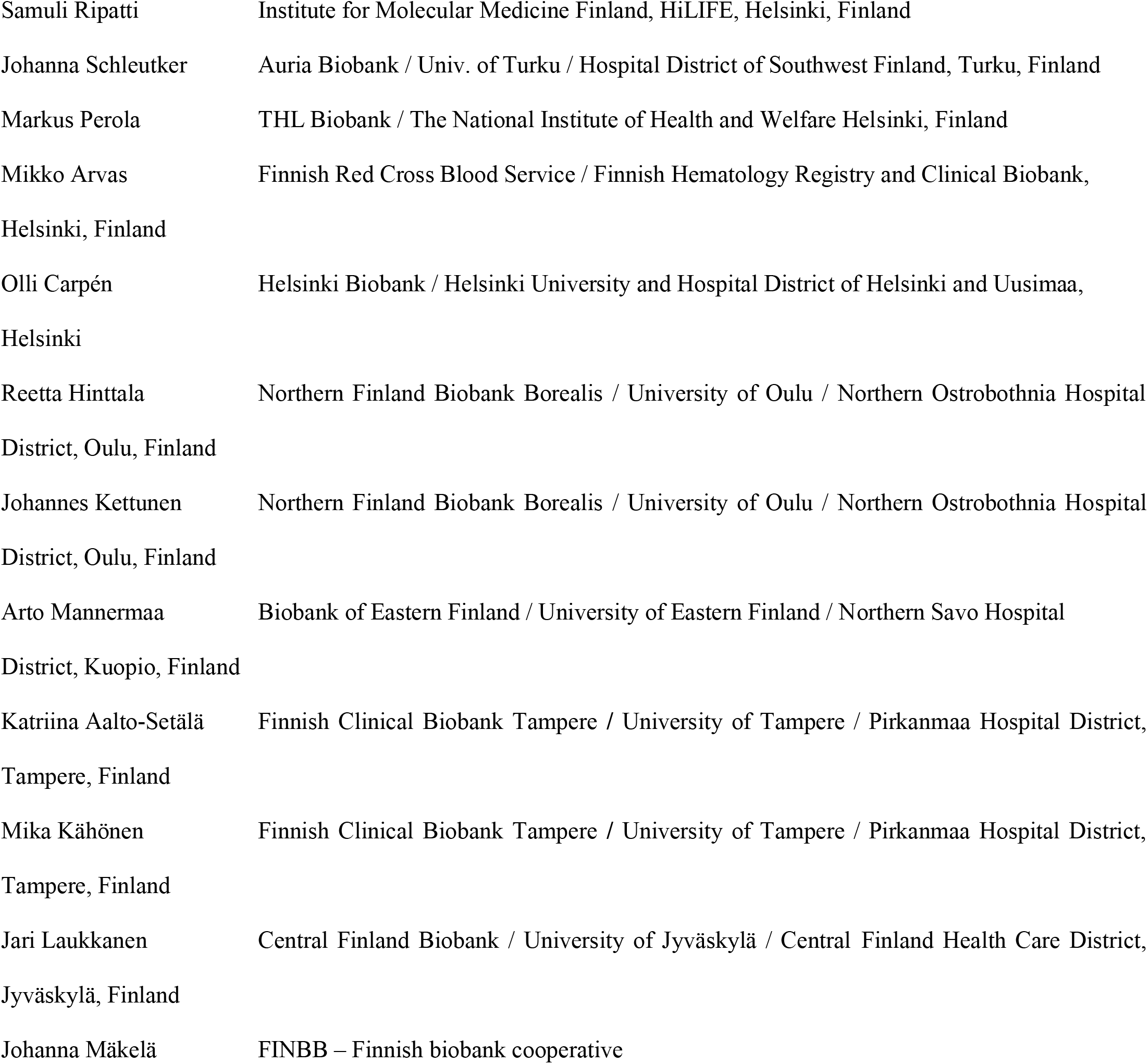

#### Clinical Groups

##### Neurology Group

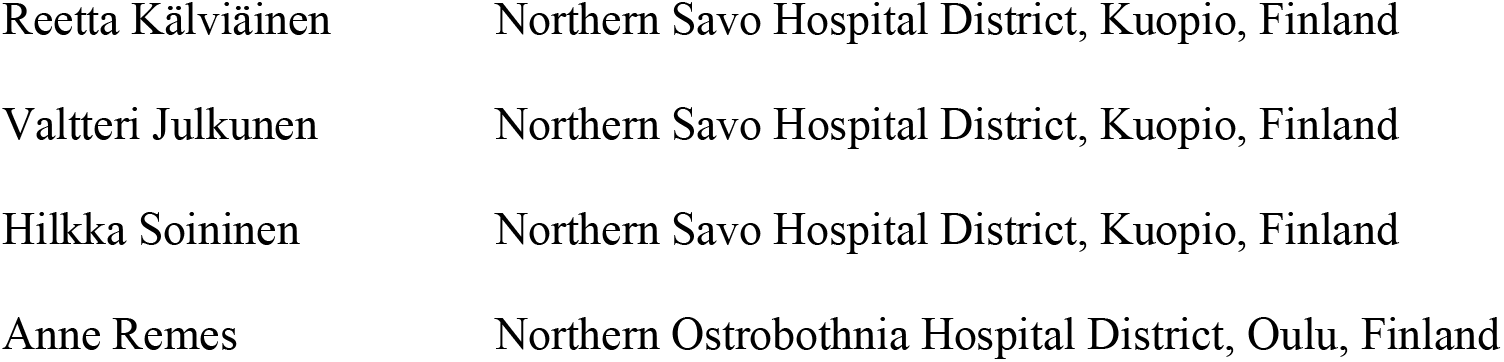
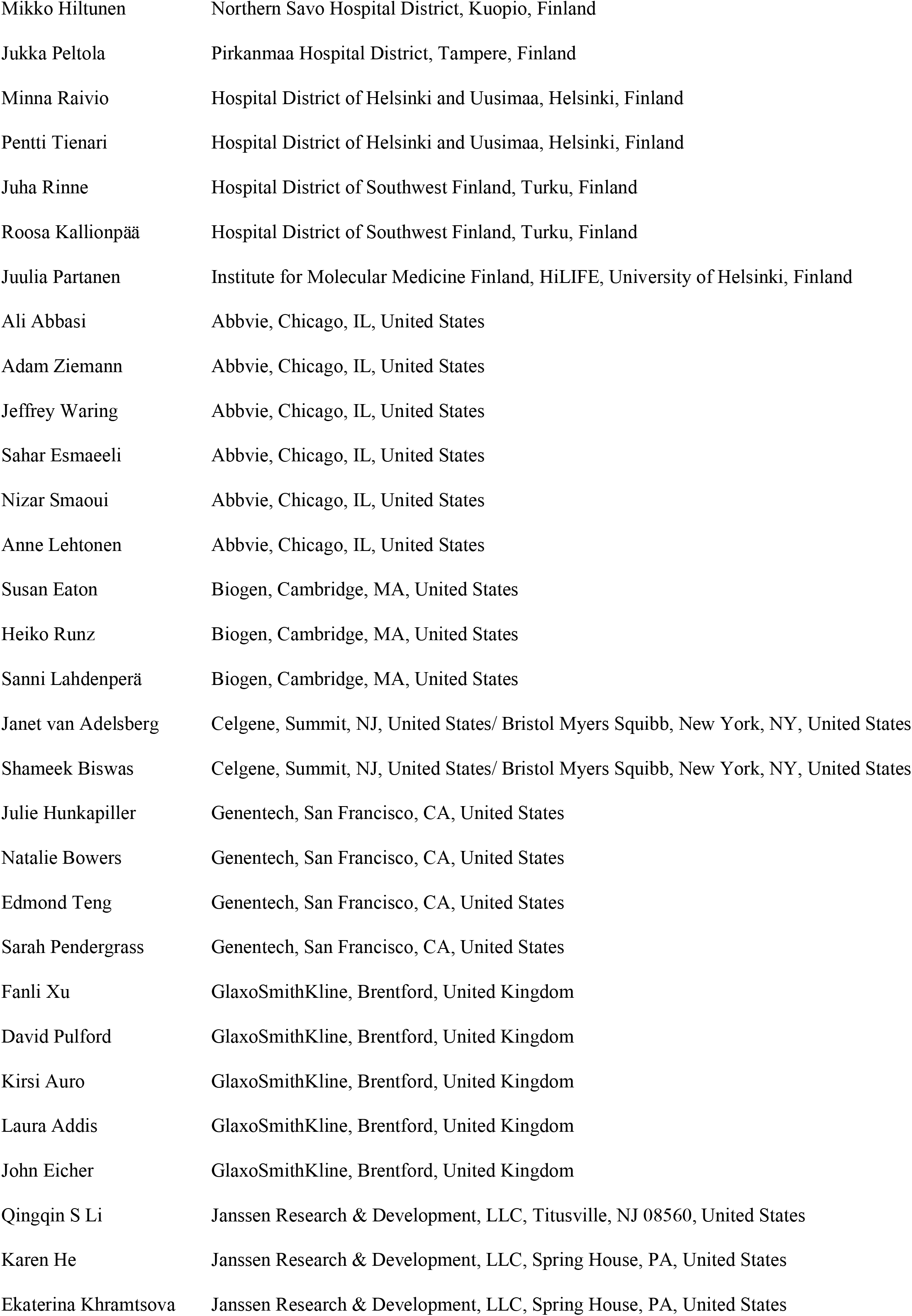
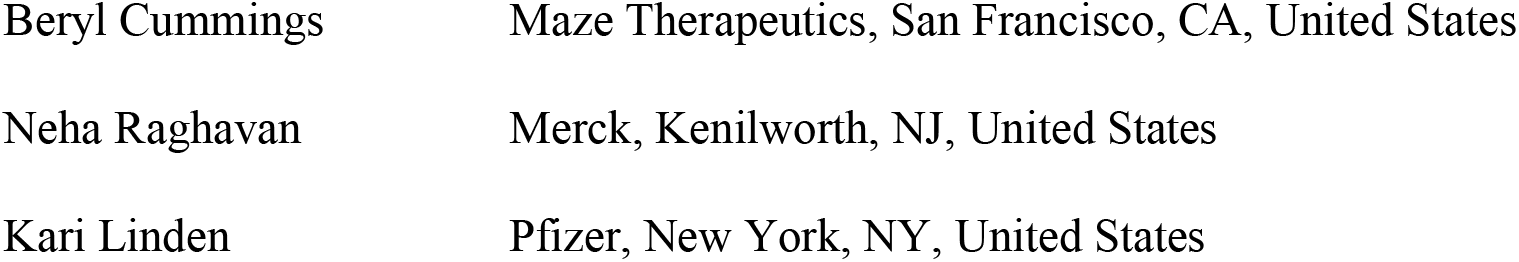

##### Gastroenterology Group

|  |  |
| --- | --- |
| Martti Färkkilä | Hospital District of Helsinki and Uusimaa, Helsinki, Finland |
| Jukka Koskela | Hospital District of Helsinki and Uusimaa, Helsinki, Finland |
| Sampsa Pikkarainen | Hospital District of Helsinki and Uusimaa, Helsinki, Finland |
| Airi Jussila | Pirkanmaa Hospital District, Tampere, Finland |
| Katri Kaukinen | Pirkanmaa Hospital District, Tampere, Finland |
| Timo Blomster | Northern Ostrobothnia Hospital District, Oulu, Finland |
| Mikko Kiviniemi | Northern Savo Hospital District, Kuopio, Finland |
| Markku Voutilainen | Hospital District of Southwest Finland, Turku, Finland |
| Mark Daly | Institute for Molecular Medicine Finland, HiLIFE, University of Helsinki, Finland |
| Ali Abbasi | Abbvie, Chicago, IL, United States |
| Graham Heap | Abbvie, Chicago, IL, United States |
| Jeffrey Waring | Abbvie, Chicago, IL, United States |
| Nizar Smaoui | Abbvie, Chicago, IL, United States |
| Fedik Rahimov | Abbvie, Chicago, IL, United States |
| Anne Lehtonen | Abbvie, Chicago, IL, United States |
| Keith Usiskin | Celgene, Summit, NJ, United States/ Bristol Myers Squibb, New York, NY, United States |
| Tim Lu | Genentech, San Francisco, CA, United States |
| Natalie Bowers | Genentech, San Francisco, CA, United States |
| Danny Oh | Genentech, San Francisco, CA, United States |
| Sarah Pendergrass | Genentech, San Francisco, CA, United States |
| Linda McCarthy | GlaxoSmithKline, Brentford, United Kingdom |
| Amy Hart | Janssen Research & Development, LLC, Spring House, PA, United States |
| Meijian Guan | Janssen Research & Development, LLC, Spring House, PA, United States |
| Jason Miller | Merck, Kenilworth, NJ, United States |
| Kirsi Kalpala | Pfizer, New York, NY, United States |
| Melissa Miller | Pfizer, New York, NY, United States |
| Xinli Hu | Pfizer, New York, NY, United States |

##### Rheumatology Group

|  |  |
| --- | --- |
| Kari Eklund | Hospital District of Helsinki and Uusimaa, Helsinki, Finland |
| Antti Palomäki | Hospital District of Southwest Finland, Turku, Finland |
| Pia Isomäki | Pirkanmaa Hospital District, Tampere, Finland |
| Laura Pirilä | Hospital District of Southwest Finland, Turku, Finland |
| Oili Kaipiainen-Seppänen | Northern Savo Hospital District, Kuopio, Finland |
| Johanna Huhtakangas | Northern Ostrobothnia Hospital District, Oulu, Finland |
| Nina Mars | Institute for Molecular Medicine Finland, HiLIFE, Helsinki, Finland |
| Ali Abbasi | Abbvie, Chicago, IL, United States |
| Jeffrey Waring | Abbvie, Chicago, IL, United States |
| Fedik Rahimov | Abbvie, Chicago, IL, United States |
| Apinya Lertratanakul | Abbvie, Chicago, IL, United States |
| Nizar Smaoui | Abbvie, Chicago, IL, United States |
| Anne Lehtonen | Abbvie, Chicago, IL, United States |
| David Close | Astra Zeneca, Cambridge, United Kingdom |
| Marla Hochfeld | Celgene, Summit, NJ, United States/ Bristol Myers Squibb, New York, NY, United States |
| Natalie Bowers | Genentech, San Francisco, CA, United States |
| Sarah Pendergrass | Genentech, San Francisco, CA, United States |
| Jorge Esparza Gordillo | GlaxoSmithKline, Brentford, United Kingdom |
| Kirsi Auro | GlaxoSmithKline, Brentford, United Kingdom |
| Dawn Waterworth | Janssen Research & Development, LLC, Spring House, PA, United States |
| Fabiana Farias | Merck, Kenilworth, NJ, United States |
| Kirsi Kalpala | Pfizer, New York, NY, United States |
| Nan Bing | Pfizer, New York, NY, United States |
| Xinli Hu | Pfizer, New York, NY, United States |

##### Pulmonology Group

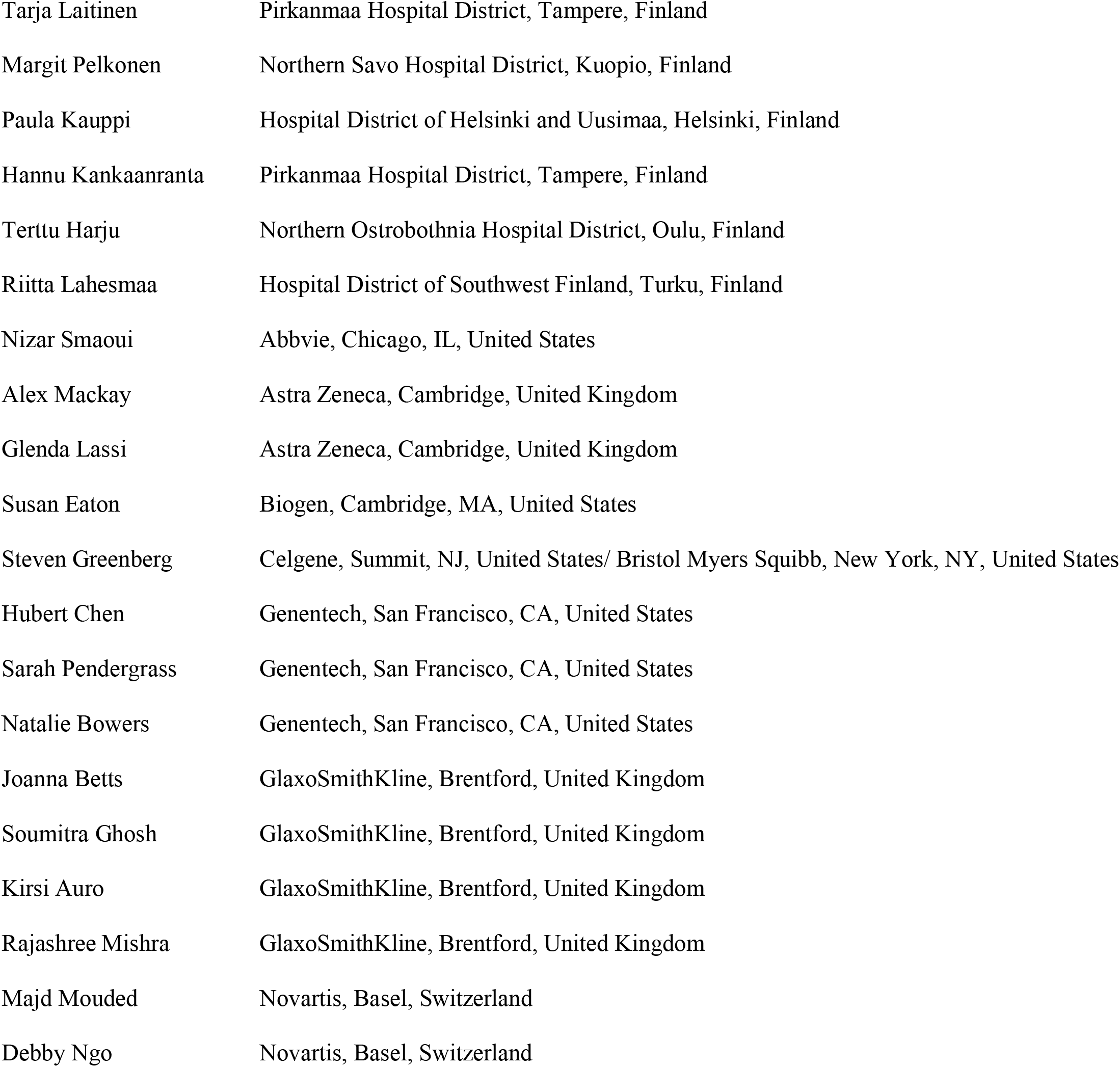

##### Cardiometabolic Diseases Group

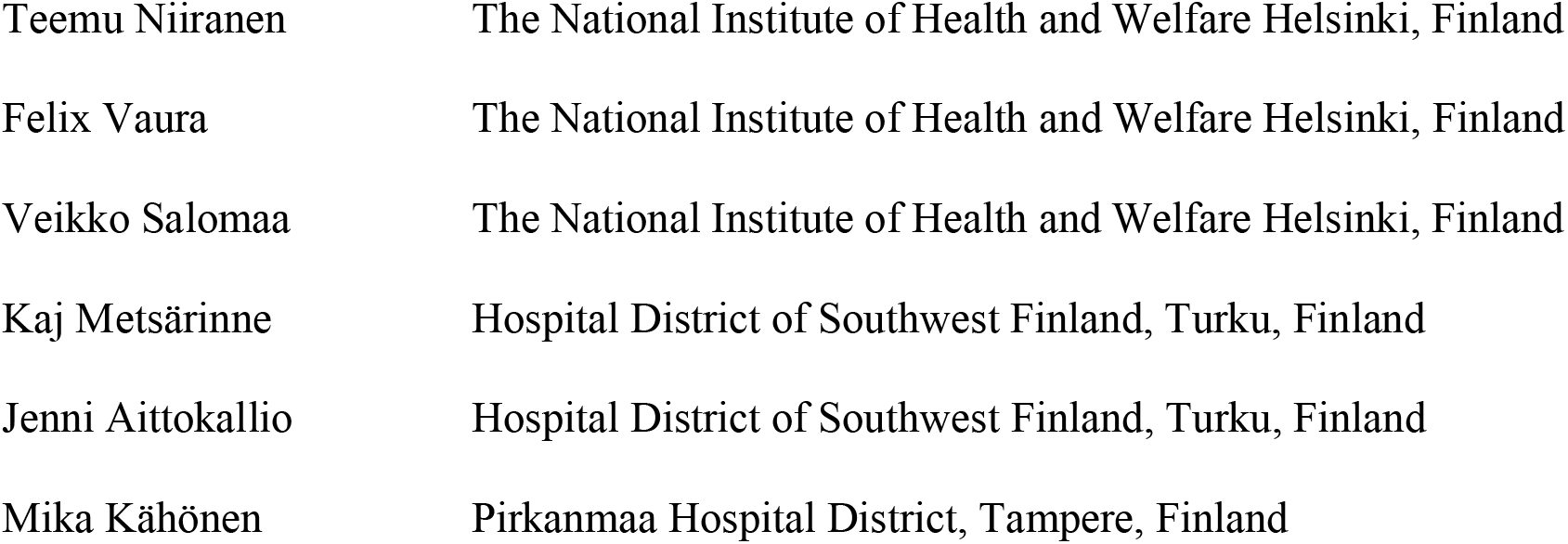
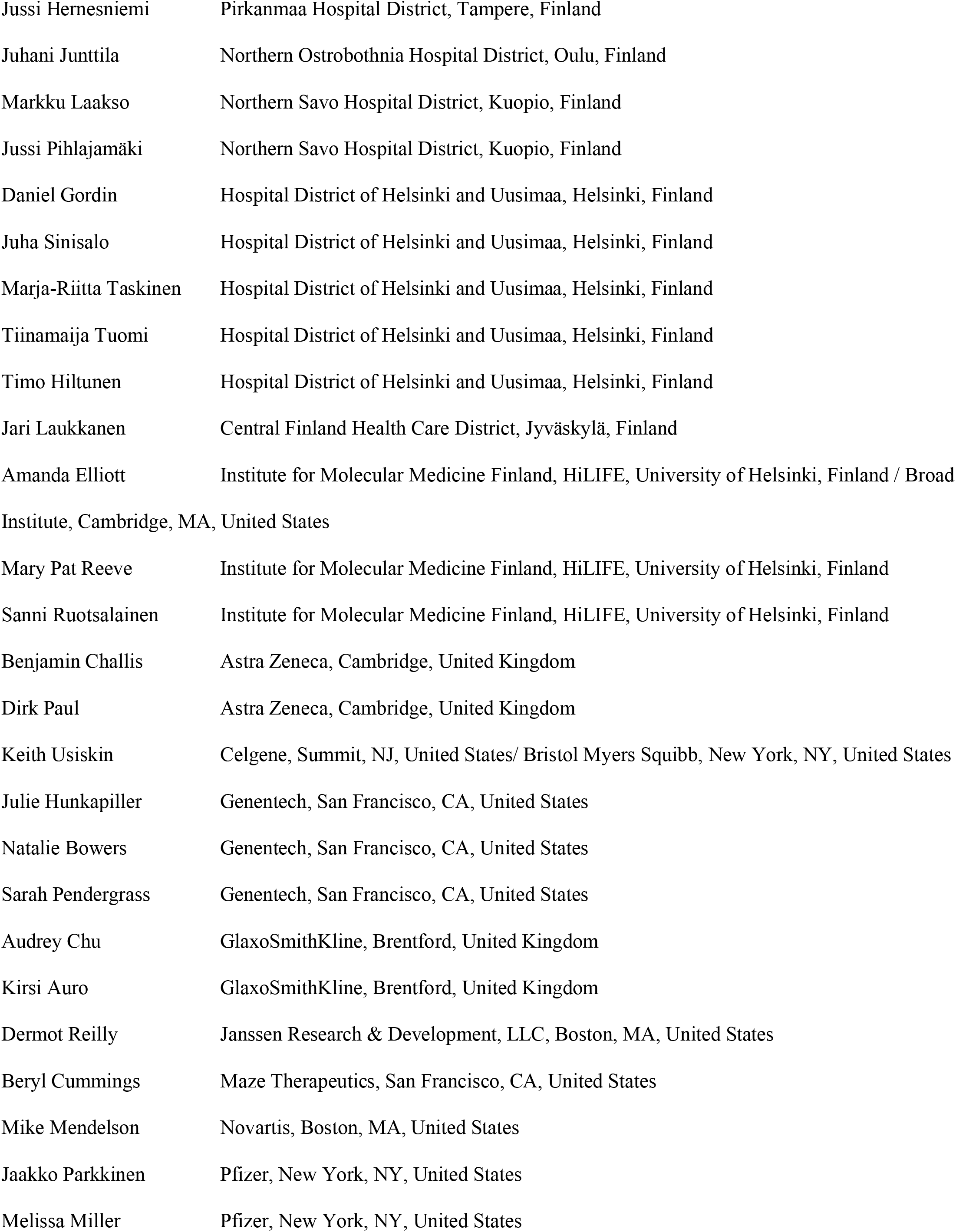

##### Oncology Group

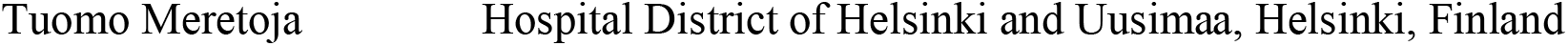
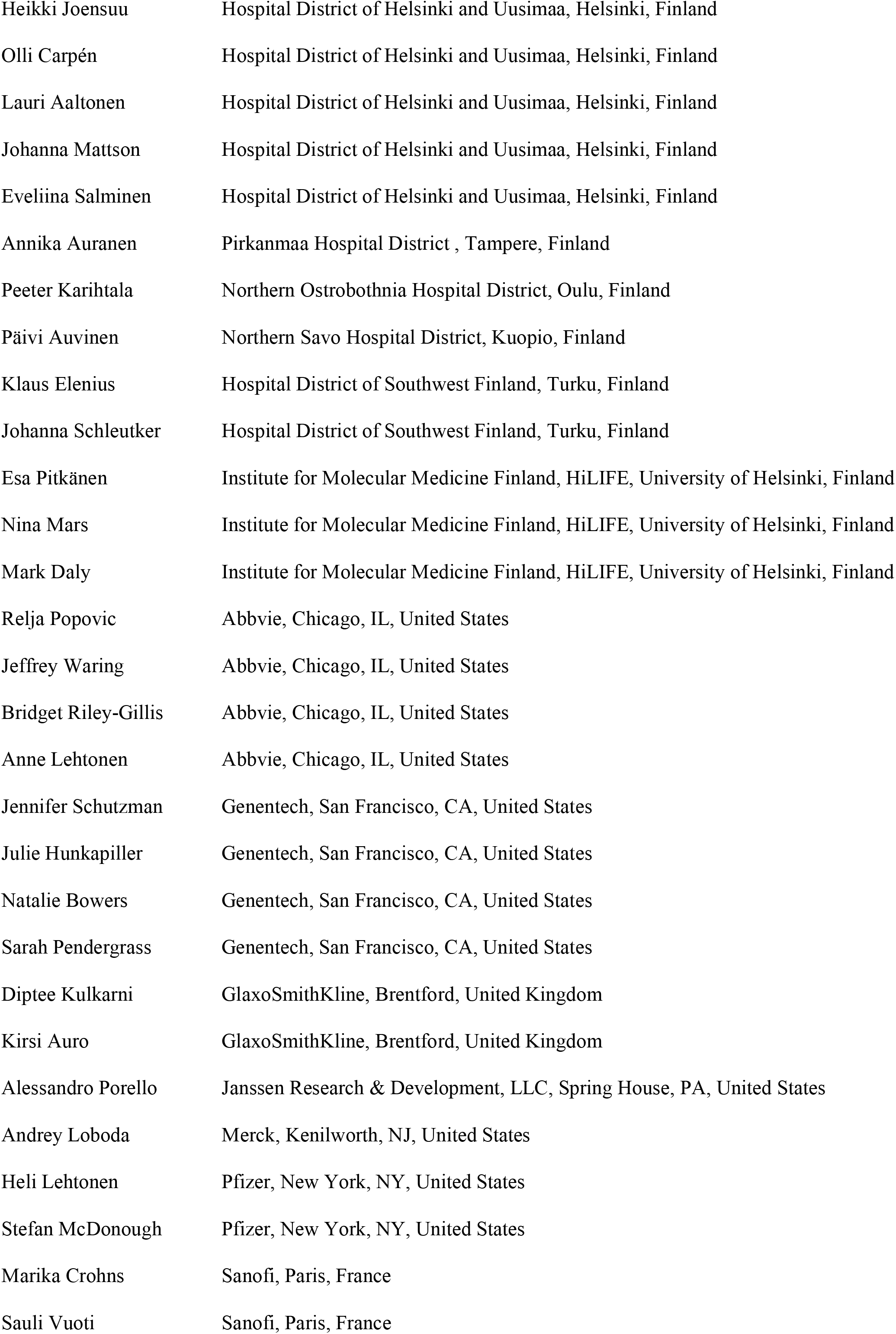

##### Opthalmology Group

**Ophthalmology Group**
|  |  |
| --- | --- |
| Kai Kaarniranta | Northern Savo Hospital District, Kuopio, Finland |
| Joni A Turunen | Hospital District of Helsinki and Uusimaa, Helsinki, Finland |
| Terhi Ollila | Hospital District of Helsinki and Uusimaa, Helsinki, Finland |
| Hannu Uusitalo | Pirkanmaa Hospital District, Tampere, Finland |
| Juha Karjalainen | Institute for Molecular Medicine Finland, HiLIFE, University of Helsinki, Finland |
| Esa Pitkänen | Institute for Molecular Medicine Finland, HiLIFE, University of Helsinki, Finland |
| Mengzhen Liu | Abbvie, Chicago, IL, United States |
| Heiko Runz | Biogen, Cambridge, MA, United States |
| Stephanie Loomis | Biogen, Cambridge, MA, United States |
| Erich Strauss | Genentech, San Francisco, CA, United States |
| Natalie Bowers | Genentech, San Francisco, CA, United States |
| Hao Chen | Genentech, San Francisco, CA, United States |
| Sarah Pendergrass | Genentech, San Francisco, CA, United States |

##### Dermatology Group

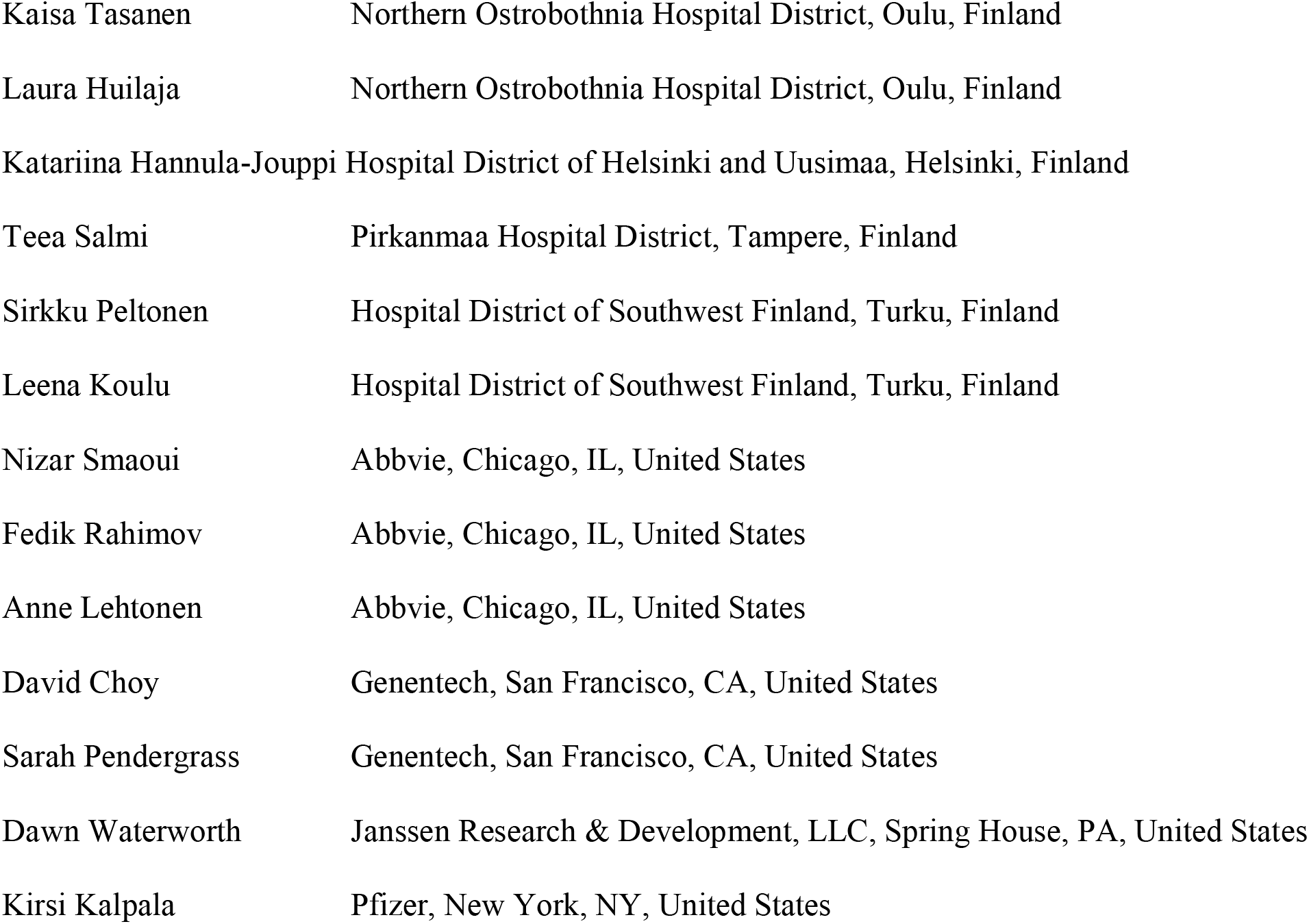
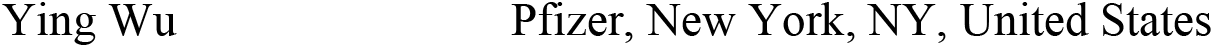

##### Odontology Group

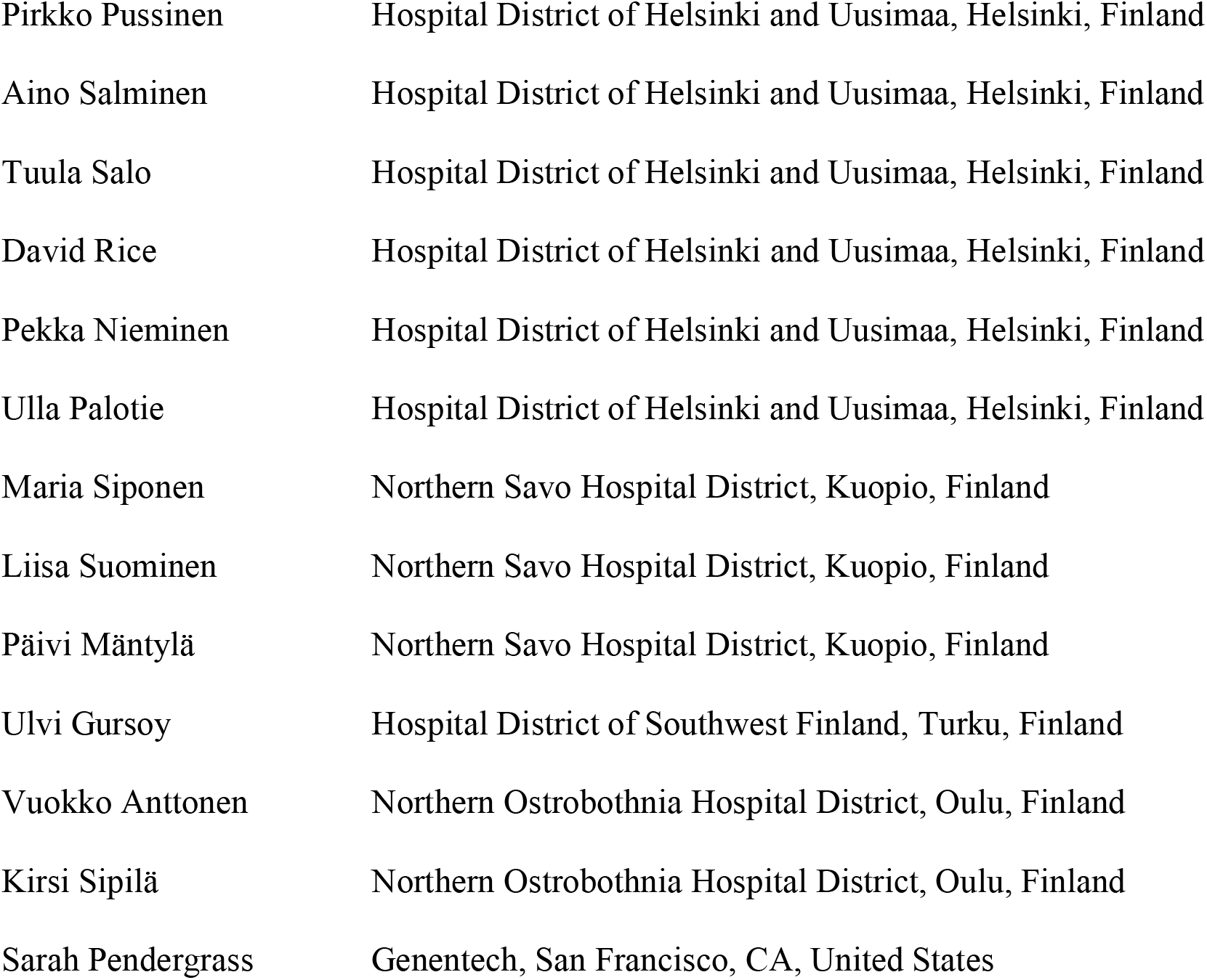

##### Women’s Health and Reproduction Group

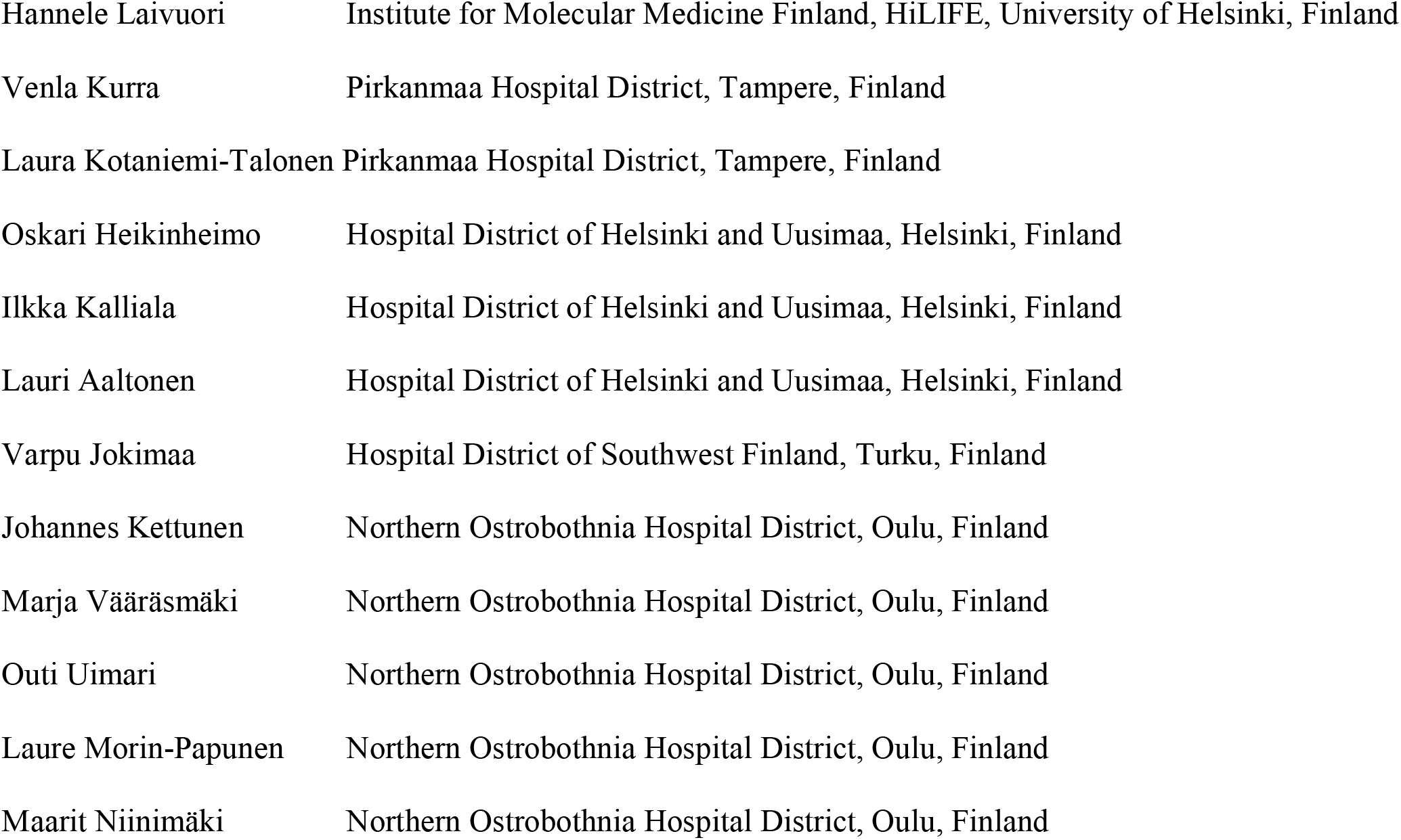
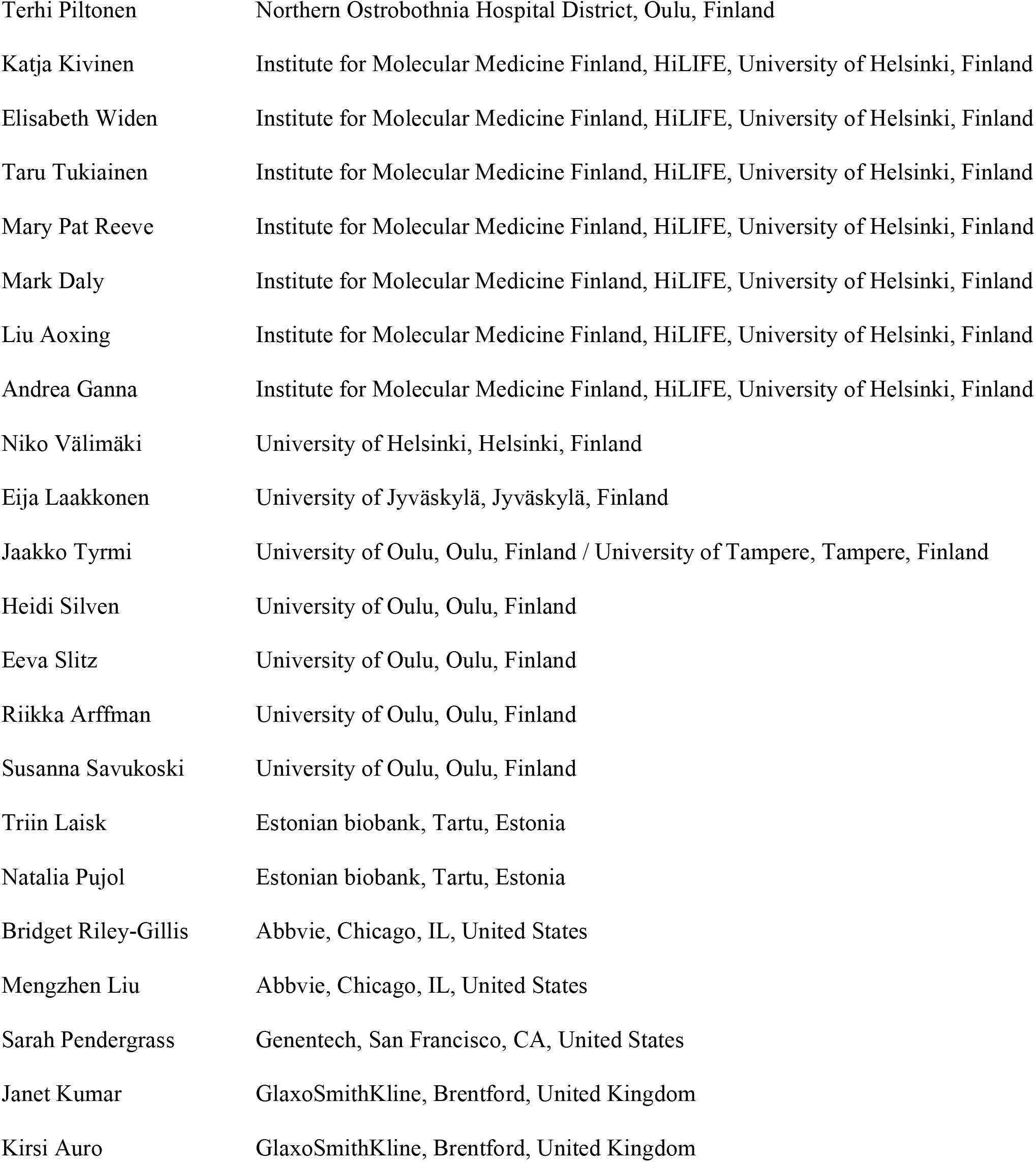

#### FinnGen Analysis working group

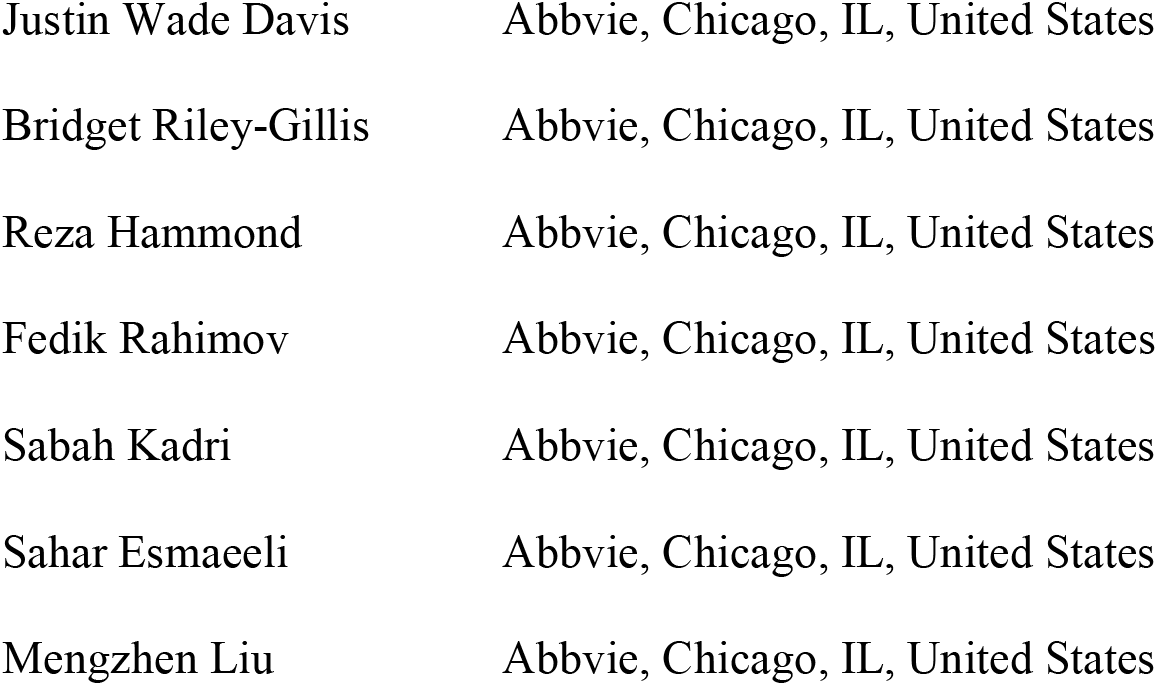
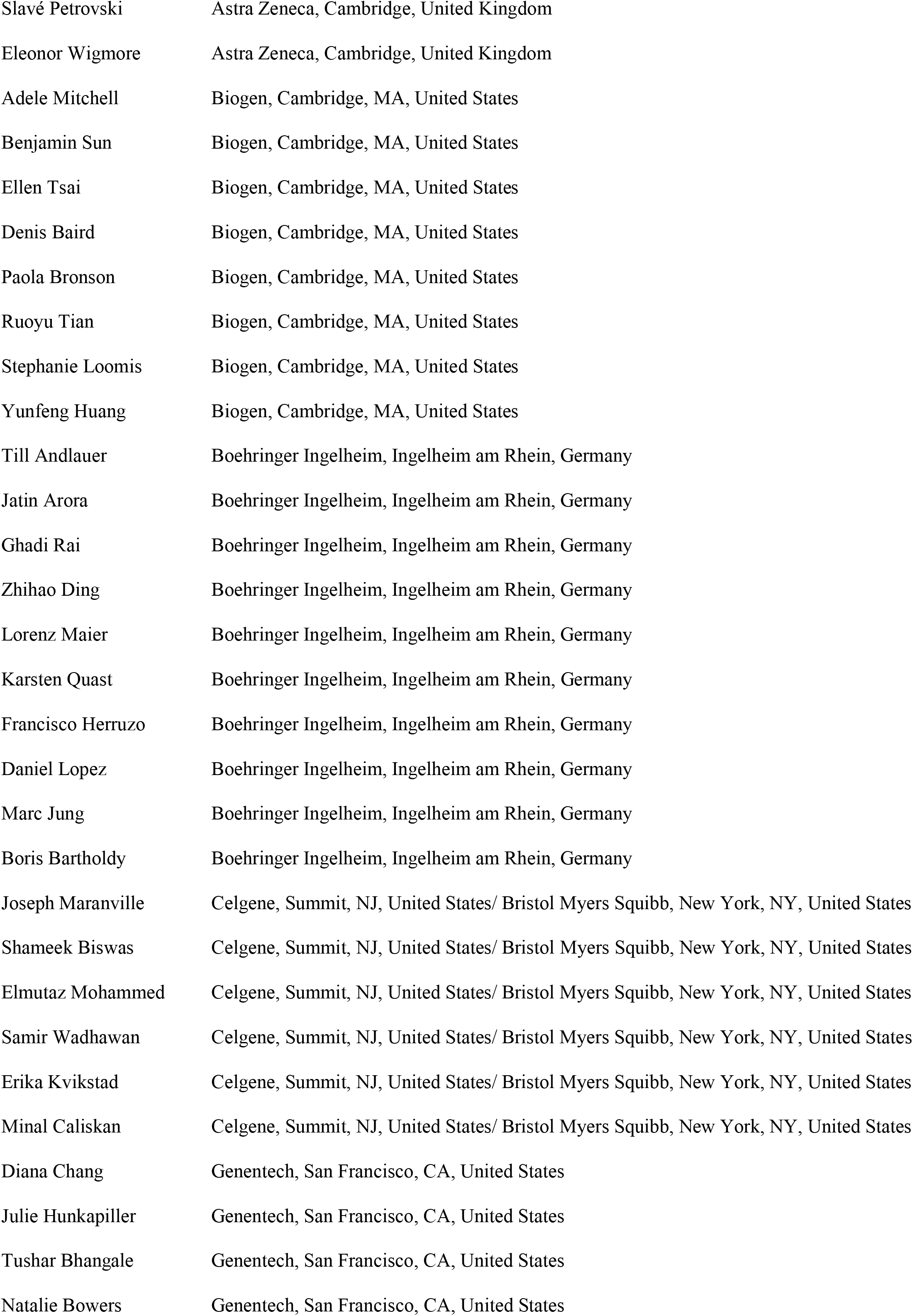
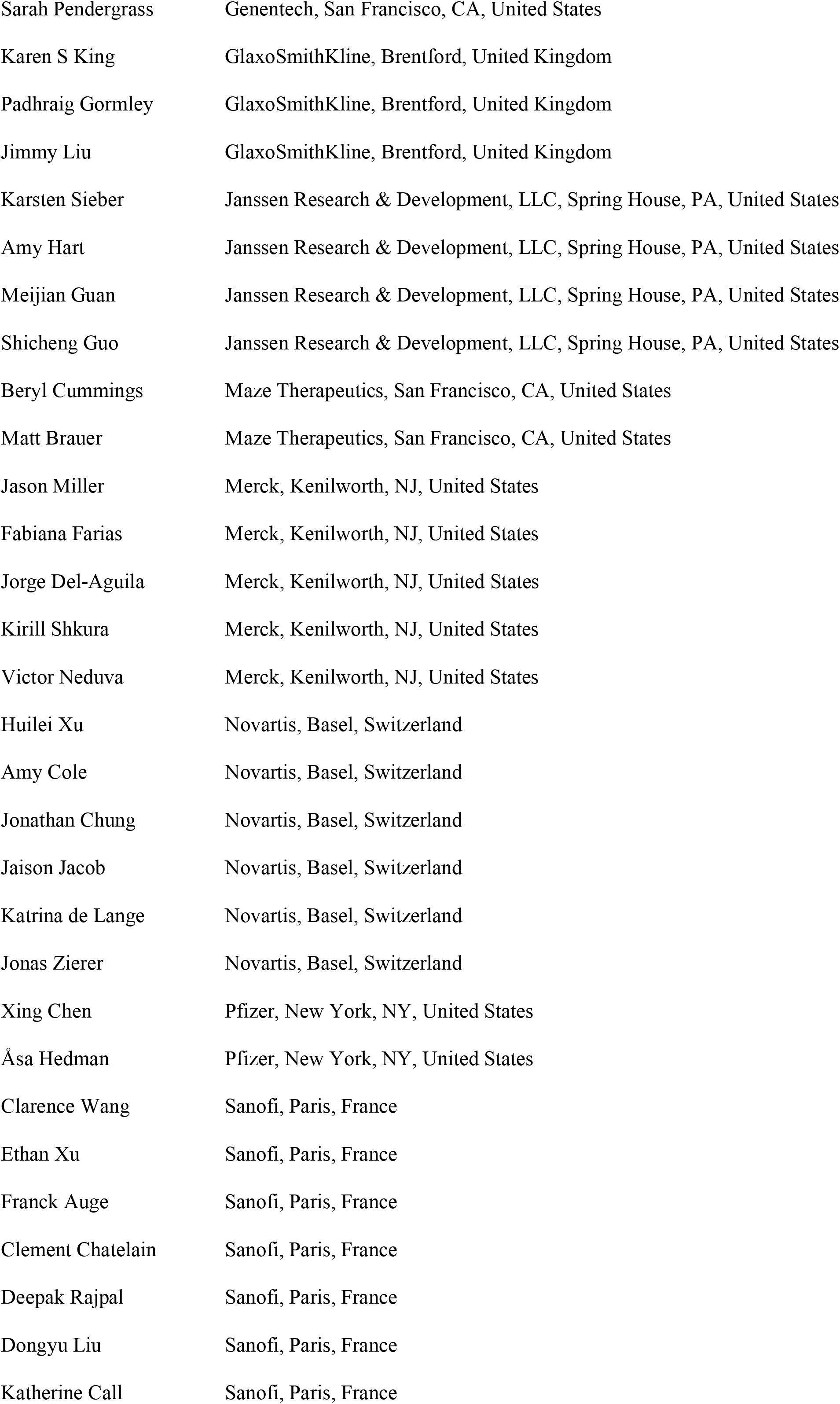
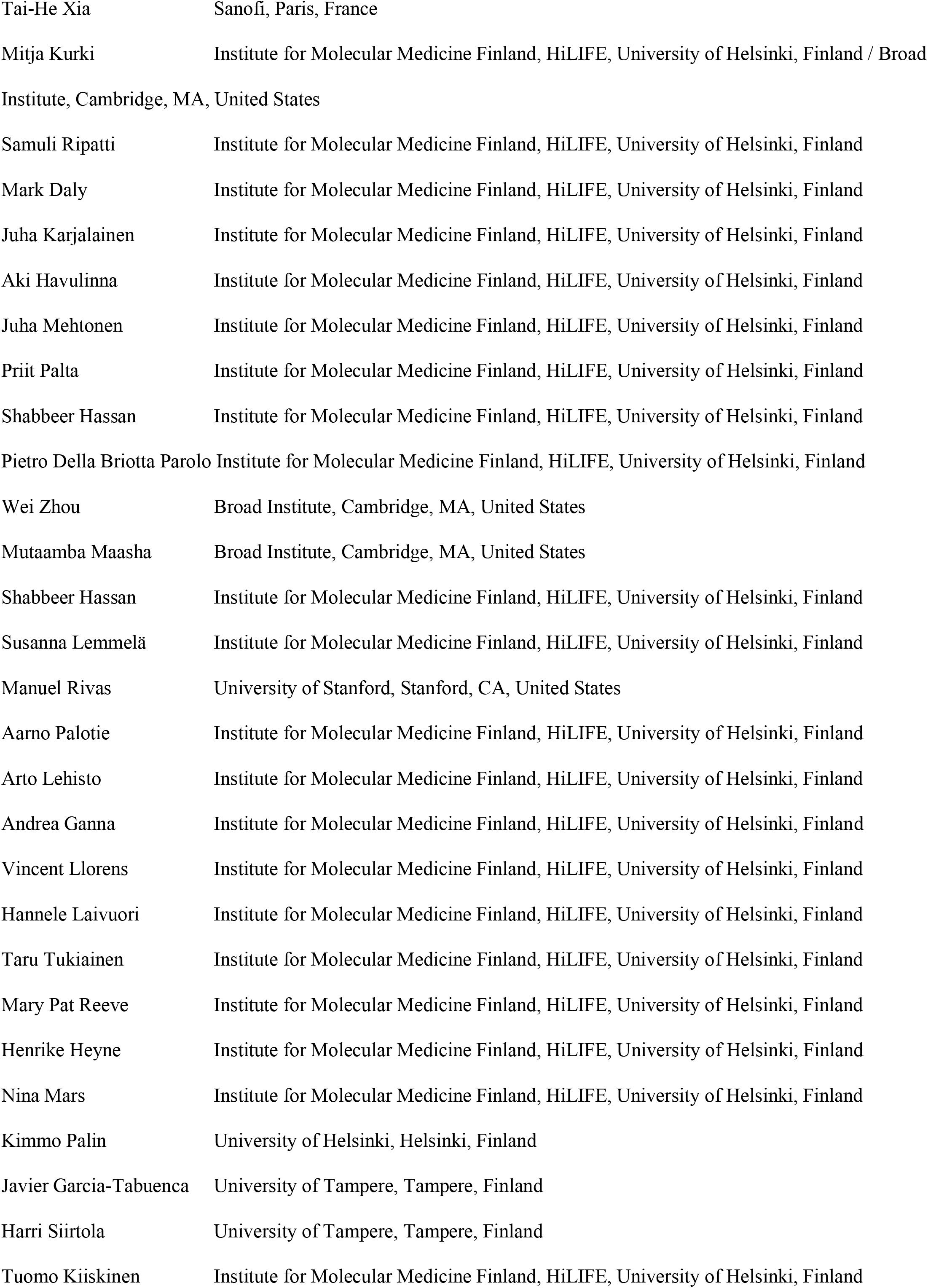
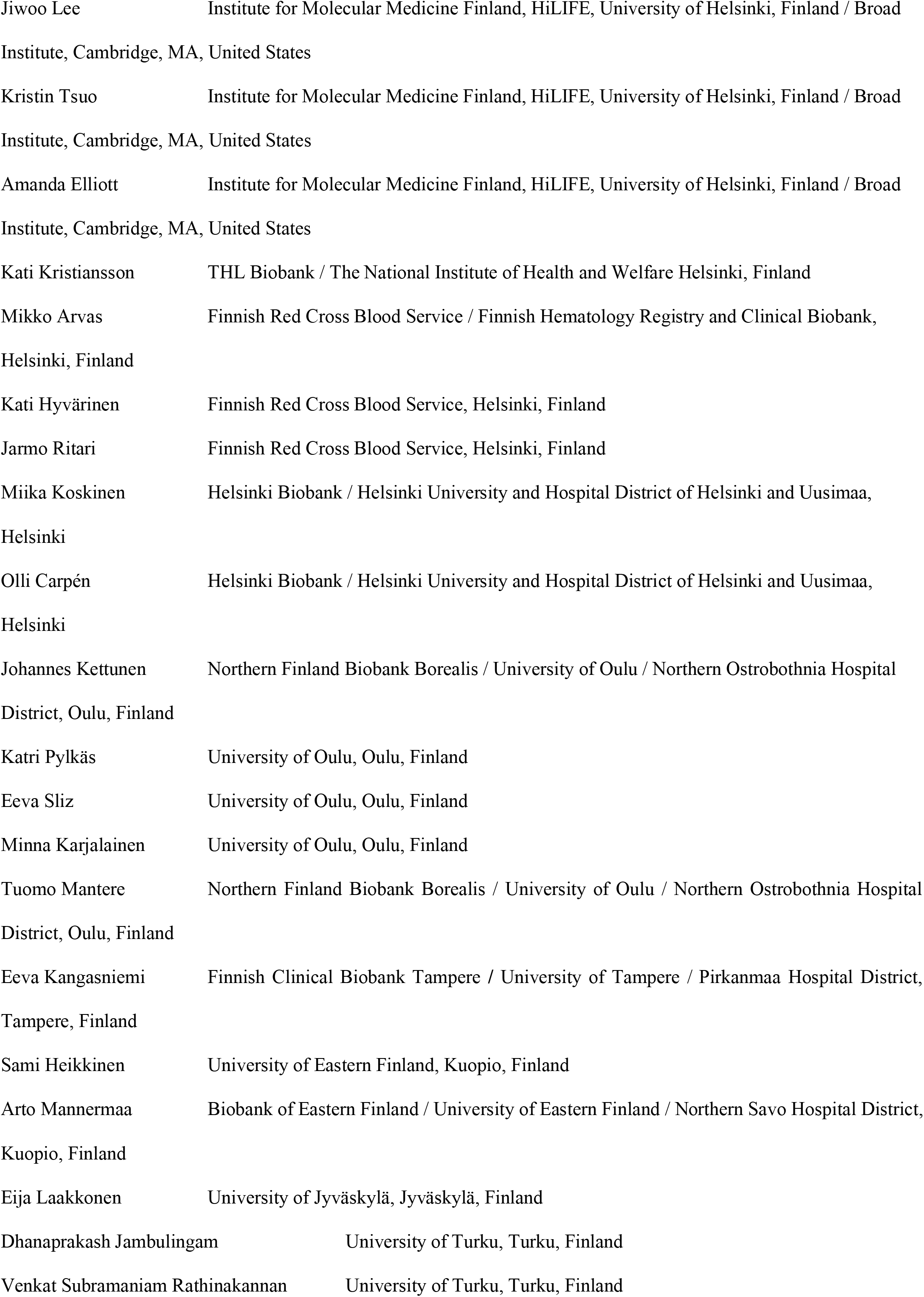
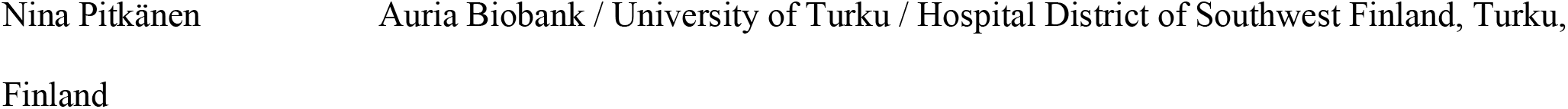

#### Biobank directors

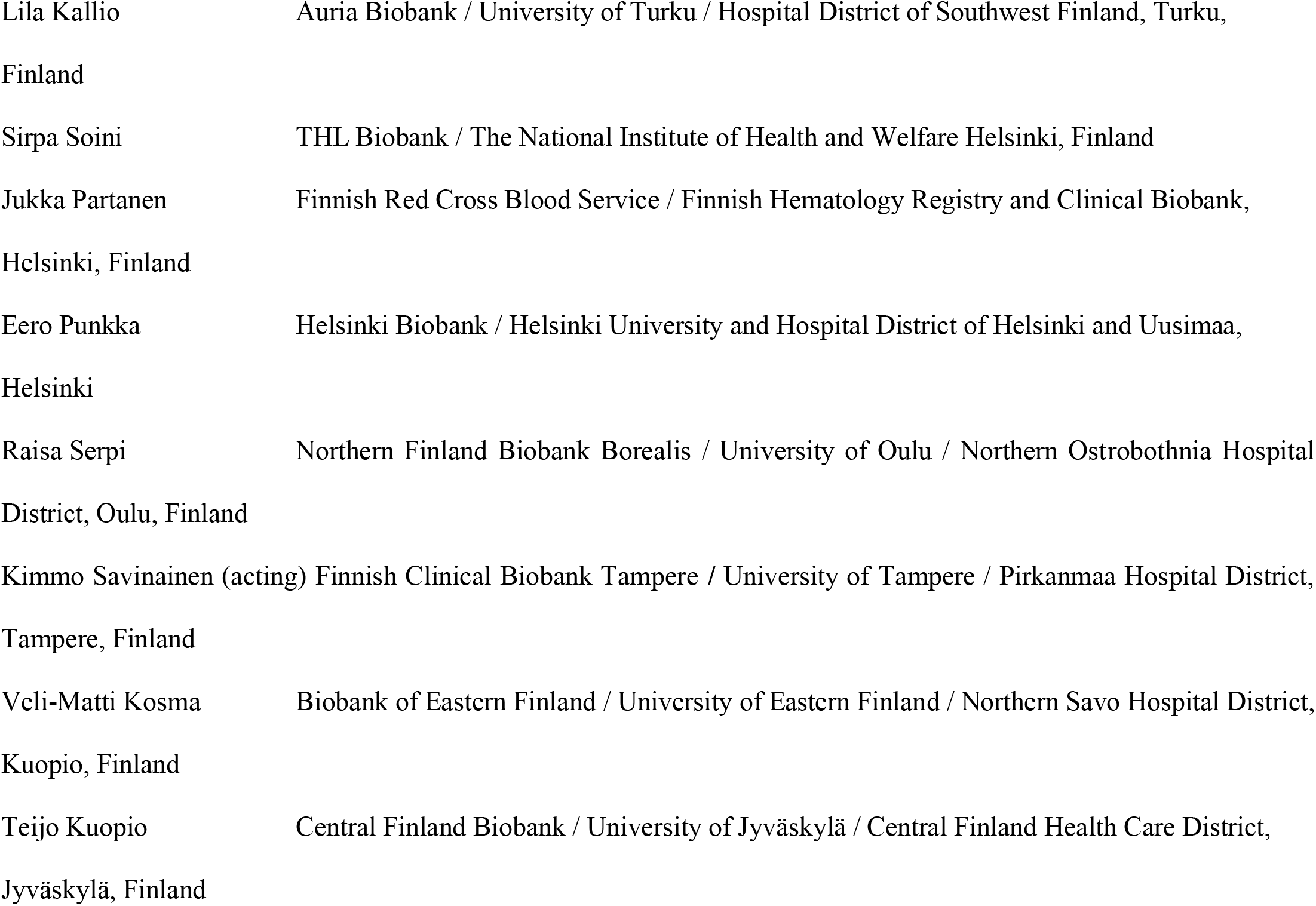

#### FinnGen Teams

##### Administration

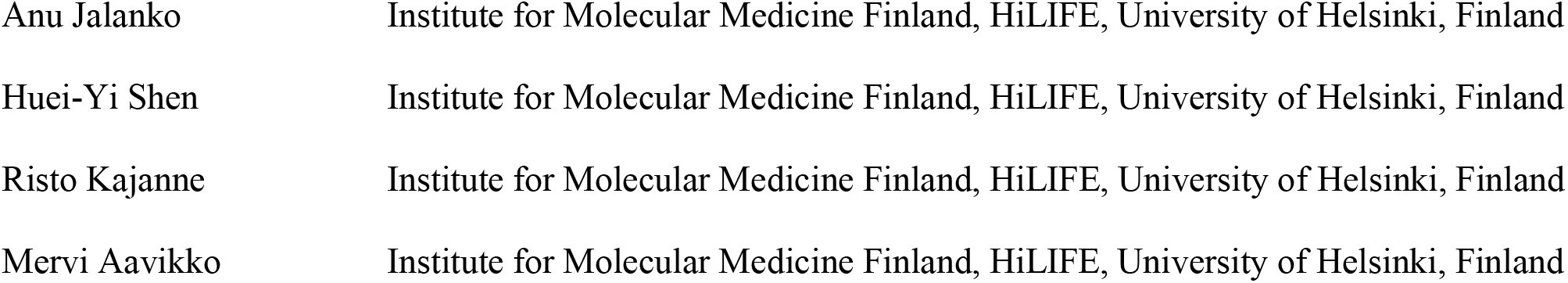

##### Analysis

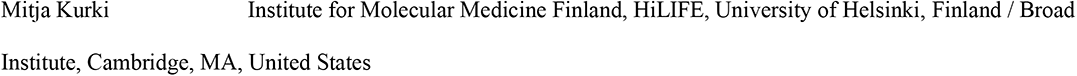
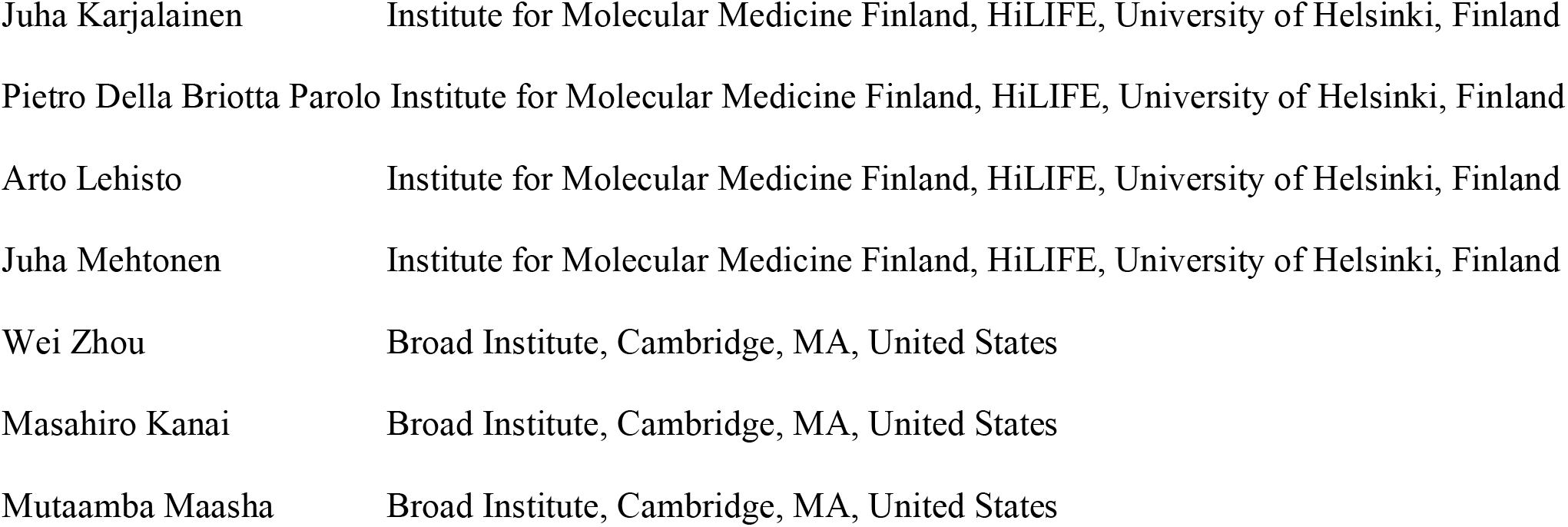

##### Clinical Endpoint Development

|  |  |
| --- | --- |
| Hannele Laivuori | Institute for Molecular Medicine Finland, HiLIFE, University of Helsinki, Finland |
| Aki Havulinna | Institute for Molecular Medicine Finland, HiLIFE, University of Helsinki, Finland |
| Susanna Lemmelä | Institute for Molecular Medicine Finland, HiLIFE, University of Helsinki, Finland |
| Tuomo Kiiskinen | Institute for Molecular Medicine Finland, HiLIFE, University of Helsinki, Finland |
| L. Elisa Lahtela | Institute for Molecular Medicine Finland, HiLIFE, University of Helsinki, Finland |

##### Communication

|  |  |
| --- | --- |
| Mari Kaunisto | Institute for Molecular Medicine Finland, HiLIFE, University of Helsinki, Finland |

##### E-Science

|  |  |
| --- | --- |
| Elina Kilpeläinen | Institute for Molecular Medicine Finland, HiLIFE, University of Helsinki, Finland |
| Timo P. Sipilä | Institute for Molecular Medicine Finland, HiLIFE, University of Helsinki, Finland |
| Oluwaseun Alexander Dada | Institute for Molecular Medicine Finland, HiLIFE, University of Helsinki, Finland |
| Awaisa Ghazal | Institute for Molecular Medicine Finland, HiLIFE, University of Helsinki, Finland |
| Anastasia Shcherban | Institute for Molecular Medicine Finland, HiLIFE, University of Helsinki, Finland |

##### Genotyping

|  |  |
| --- | --- |
| Kati Donner | Institute for Molecular Medicine Finland, HiLIFE, University of Helsinki, Finland |
| Timo P. Sipilä | Institute for Molecular Medicine Finland, HiLIFE, University of Helsinki, Finland |

##### Sample Collection Coordination

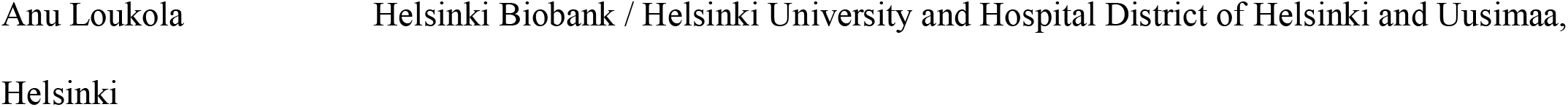

##### Sample Logistics

|  |  |
| --- | --- |
| Päivi Laiho | THL Biobank / The National Institute of Health and Welfare Helsinki, Finland |
| Tuuli Sistonen | THL Biobank / The National Institute of Health and Welfare Helsinki, Finland |
| Essi Kaiharju | THL Biobank / The National Institute of Health and Welfare Helsinki, Finland |
| Markku Laukkanen | THL Biobank / The National Institute of Health and Welfare Helsinki, Finland |
| Elina Järvensivu | THL Biobank / The National Institute of Health and Welfare Helsinki, Finland |
| Sini Lähteenmäki | THL Biobank / The National Institute of Health and Welfare Helsinki, Finland |
| Lotta Männikkö | THL Biobank / The National Institute of Health and Welfare Helsinki, Finland |
| Regis Wong | THL Biobank / The National Institute of Health and Welfare Helsinki, Finland |

##### Registry Data Operations

|  |  |
| --- | --- |
| Hannele Mattsson | THL Biobank / The National Institute of Health and Welfare Helsinki, Finland |
| Kati Kristiansson | THL Biobank / The National Institute of Health and Welfare Helsinki, Finland |
| Susanna Lemmelä | Institute for Molecular Medicine Finland, HiLIFE, University of Helsinki, Finland |
| Sami Koskelainen | THL Biobank / The National Institute of Health and Welfare Helsinki, Finland |
| Tero Hiekkalinna | THL Biobank / The National Institute of Health and Welfare Helsinki, Finland |
| Teemu Paajanen | THL Biobank / The National Institute of Health and Welfare Helsinki, Finland |

##### Sequencing Informatics

|  |  |
| --- | --- |
| Priit Palta | Institute for Molecular Medicine Finland, HiLIFE, University of Helsinki, Finland |
| Kalle Pärn | Institute for Molecular Medicine Finland, HiLIFE, University of Helsinki, Finland |
| Mart Kals | Institute for Molecular Medicine Finland, HiLIFE, University of Helsinki, Finland |
| Shuang Luo | Institute for Molecular Medicine Finland, HiLIFE, University of Helsinki, Finland |
| Vishal Sinha | Institute for Molecular Medicine Finland, HiLIFE, University of Helsinki, Finland |

##### Trajectory

|  |  |
| --- | --- |
| Tarja Laitinen | Pirkanmaa Hospital District, Tampere, Finland |
| Mary Pat Reeve | Institute for Molecular Medicine Finland, HiLIFE, University of Helsinki, Finland |
| Marianna Niemi | University of Tampere, Tampere, Finland |
| Harri Siirtola | University of Tampere, Tampere, Finland |
| Javier Gracia-Tabuenca | University of Tampere, Tampere, Finland |
| Mika Helminen | University of Tampere, Tampere, Finland |
| Tiina Luukkaala | University of Tampere, Tampere, Finland |
| Iida Vähätalo | University of Tampere, Tampere, Finland |

##### Data protection officer

|  |  |
| --- | --- |
| Tero Jyrhämä | Institute for Molecular Medicine Finland, HiLIFE, University of Helsinki, Finland |

##### FINBB – Finnish biobank cooperative

Marco Hautalahti

Johanna Mäkelä

Laura Mustaniemi

Mirkka Koivusalo

Sarah Smith

Tom Southerington

