## Supplemental Table 1-15 for "Integration of biomarker polygenic risk score improves prediction of coronary heart disease in UK Biobank and FinnGen"

Table S1: PRS HR and variance explained of selected optimal biomarkers

| CHD Optimal Biomarkers | PRS HR (CI) for CHD per 1SD | PRS HR (CI) for CHD per 1SD | PRS HR (CI) for CHD per 1SD | PRS HR (CI) for CHD per 1SD | PRS HR (CI) for CHD per 1SD | PRS HR (CI) for CHD per 1SD |
|---|---|---|---|---|---|---|
| | UKB Test | UKB Test Female | UKB Test Male | FinnGen | FinnGen Female | FinnGen Male |
| **Apo-A1** | 0.89 (0.83-0.95) | 0.88 (0.77-1.00) | 0.89 (0.83-0.96) | 0.91 (0.90-0.92) | 0.92 (0.90-0.94) | 0.91 (0.90-0.92) |
| **ApoB*** | 1.32 (1.24-1.41) | 1.33 (1.16-1.52) | 1.32 (1.23-1.42) | 1.18 (1.17-1.20) | 1.14 (1.12-1.17) | 1.20 (1.18-1.22) |
| **CPD*** | 1.12 (1.05-1.19) | 1.14 (1.00-1.30) | 1.11 (1.03-1.19) | 1.05 (1.03-1.06) | 1.03 (1.01-1.06) | 1.05 (1.04-1.07) |
| **CREA** | 1.00 (0.93-1.06) | 0.93 (0.81-1.06) | 1.02 (0.95-1.09) | 0.96 (0.95-0.97) | 0.97 (0.95-1.00) | 0.96 (0.95-0.97) |
| **C-Reactive Protein*** | 1.02 (0.96-1.08) | 1.07 (0.93-1.22) | 1.00 (0.93-1.08) | 1.01 (0.99-1.02) | 1.02 (1.00-1.05) | 1.00 (0.99-1.02) |
| **HbA1c*** | 1.09 (1.02-1.16) | 1.01 (0.89-1.16) | 1.12 (1.04-1.20) | 1.06 (1.04-1.07) | 1.06 (1.04-1.09) | 1.05 (1.04-1.07) |
| **HDL*** | 0.87 (0.82-0.93) | 0.84 (0.74-0.96) | 0.88 (0.82-0.95) | 0.90 (0.89-0.91) | 0.91 (0.88-0.93) | 0.90 (0.89-0.92) |
| **LDL*** | 1.30 (1.22-1.38) | 1.30 (1.14-1.50) | 1.29 (1.20-1.39) | 1.17 (1.16-1.18) | 1.13 (1.11-1.15) | 1.19 (1.17-1.20) |
| **SBP*** | 1.26 (1.18-1.34) | 1.34 (1.17-1.53) | 1.23 (1.15-1.33) | 1.19 (1.17-1.20) | 1.22 (1.19-1.25) | 1.17 (1.16-1.19) |
| **TRIG*** | 1.19 (1.12-1.27) | 1.29 (1.13-1.47) | 1.16 (1.08-1.25) | 1.09 (1.08-1.10) | 1.09 (1.07-1.10) | 1.09 (1.08-1.11) |

* Selected in elastic.net sex specific optimal models

Adjusted for age, gender and PCs1-10

| Variances explained (R-squared) UKB Test | Variances explained (R-squared) UKB Test Female | Variances explained (R-squared) UKB Test Male | PRS HR (CI) for CHD per 1SD UKB Validation |
|---|---|---|---|
| 0.097 | 0.104 | 0.136 | 0.81 (0.79-0.84) |
| 0.165 | 0.170 | 0.161 | 1.24 (1.21-1.28) |
| 0.009 | 0.008 | 0.01 | 1.11 (1.08-1.14) |
| 0.081 | 0.129 | 0.099 | 0.97 (0.94-0.99) |
| 0.106 | 0.120 | 0.089 | 1.01 (0.98-1.04) |
| 0.129 | 0.129 | 0.130 | 1.10 (1.07-1.13) |
| 0.129 | 0.171 | 0.156 | 0.80 (0.78-0.82) |
| 0.156 | 0.162 | 0.149 | 1.21 (1.18-1.24) |
| 0.073 | 0.077 | 0.071 | 1.26 (1.23-1.30) |
| 0.103 | 0.110 | 0.111 | 1.11 (1.08-1.15) |

Table S2: UKB Training PRSes CHD survival HR (CI)/1 SD

| PRS | Quant vs 1-Q | HR (CI) | Z | CIndex (SE) | AUC (CI) |
|---|---|---|---|---|---|
| **UKB Training** | | | | | **Baseline .759 (.754-.763)** |
| **BioPRS** | | 1.43 (1.41-1.46) | 38.43 | .763 (.002) | .775 (.771-.780) |
| | Q80+ | 1.85 (1.77-1.92) | 30.39 | .755 (.002) | .768 (.764-.772) |
| | Q90+ | 1.92 (1.82-2.01) | 26.06 | .752 (.002) | .765 (.761-.769) |
| | Q95+ | 1.96 (1.84-2.09) | 20.51 | .750 (.002) | .763 (.758-.766) |
| | Q99+ | 2.07 (1.81-2.38) | 10.45 | .747 (.002) | .760 (.755-.764) |
| **CHDPRS** | | 1.64 (1.61-1.67) | 52.66 | .776 (.002) | .787 (.783-.791) |
| | Q80+ | 2.30 (2.21-2.39) | 42.75 | .764 (.002) | .776 (.772-.780) |
| | Q90+ | 2.46 (2.35-2.57) | 38.80 | .759 (.002) | .772 (.768-.776) |
| | Q95+ | 2.70 (2.55-2.86) | 34.26 | .755 (.002) | .768 (.764-.772) |
| | Q99+ | 3.66 (3.29-4.06) | 24.23 | .750 (.002) | .762 (.758-.767) |
| **CHDBioPRS** | | 1.75 (1.72-1.78) | 59.20 | .783 (.002) | .795 (.791-.799) |
| | Q80+ | 2.56 (2.46-2.66) | 48.9 | .769 (.002) | .781 (.777-.785) |
| | Q90+ | 2.77 (2.65-2.89) | 45.29 | .763 (.002) | .776 (.771-.779) |
| | Q95+ | 2.98 (2.82-3.15) | 38.64 | .757 (.002) | .770 (.766-.774) |
| | Q99+ | 3.66 (3.30-4.07) | 24.02 | .750 (.002) | .763 (.759-.767) |

Adjusted for age, gender and PCs1-10

Table S3: UKB Validation PRSes CHD survival HR (CI)/1 SD

| PRS | Quant vs 1-Q | HR (CI) | Z | CIndex (SE) | AUC (CI) |
|---|---|---|---|---|---|
| **UKB Validation** | | | | | **Baseline .762 (.756-.768)** |
| **BioPRS** | | 1.41 (1.37-1.45) | 23.78 | .763 (.003) | .777 (.771-.783) |
| | Q80+ | 1.74 (1.64-1.85) | 17.77 | .755 (.003) | .770 (.764-.776) |
| | Q90+ | 1.82 (1.69-1.96) | 15.47 | .753 (.003) | .768 (.761-.774) |
| | Q95+ | 1.92 (1.74-2.12) | 12.84 | .750 (.003) | .765 (.759-.772) |
| | Q99+ | 2.17 (1.78-2.66) | 7.59 | .748 (.003) | .763 (.757-.770) |
| **CHDPRS** | | 1.62 (1.57-1.67) | 33.67 | .776 (.003) | .790 (.784-.796) |
| | Q80+ | 2.22 (2.09-2.35) | 26.69 | .764 (.003) | .778 (.772-.784) |
| | Q90+ | 2.43 (2.27-2.61) | 25.09 | .761 (.003) | .775 (.769-.781) |
| | Q95+ | 2.65 (2.43-2.89) | 21.74 | .757 (.003) | .772 (.765-.778) |
| | Q99+ | 3.50 (2.96-4.13) | 14.80 | .751 (.003) | .766 (.760-.772) |
| **CHDBioPRS** | | 1.72 (1.67-1.77) | 37.56 | .783 (.003) | .795 (.789-.801) |
| | Q80+ | 2.48 (2.34-2.63) | 30.95 | .769 (.003) | .783 (.777-.789) |
| | Q90+ | 2.63 (2.46-2.81) | 27.80 | .763 (.003) | .777 (.771-.784) |
| | Q95+ | 2.80 (2.57-3.06) | 23.12 | .758 (.003) | .772 (.767-.779) |
| | Q99+ | 3.98 (3.40-4.68) | 16.93 | .751 (.003) | .767 (.760-.773) |

Adjusted for age, gender and PCs1-10

Table S4: UKB Test PRSes CHD survival HR (CI)/1 SD

| PRS | Quant vs 1-Q | HR (CI) | Z | CIndex (SE) | AUC (CI) |
|---|---|---|---|---|---|
| **UKB Test** | | | | | **Baseline .777 (.763-.791)** |
| **BioPRS** | | 1.45 (1.36-1.54) | 11.27 | .786 (.007) | .794 (.780-.807) |
| | Q80+ | 1.92 (1.67-2.21) | 9.22 | .780 (.007) | .788 (.774-.802) |
| | Q90+ | 2.18 (1.85-2.57) | 9.21 | .779 (.007) | .787 (.773-.801) |
| | Q95+ | 2.33 (1.89-2.87) | 7.89 | .777 (.007) | .785 (.771-.798) |
| | Q99+ | 2.35 (1.54-3.59) | 3.96 | .773 (.007) | .781 (.767-.795) |
| **CHDPRS** | | 1.78 (1.67-1.91) | 16.61 | .800 (.007) | .808 (.795-.822) |
| | Q80+ | 2.39 (2.09-2.74) | 12.48 | .786 (.007) | .794 (.781-.808) |
| | Q90+ | 2.51 (2.14-2.95) | 11.16 | .782 (.007) | .790 (.776-.804) |
| | Q95+ | 3.14 (2.58-3.82) | 11.47 | .778 (.007) | .787 (.772-.800) |
| | Q99+ | 3.52 (2.36-5.26) | 6.14 | .774 (.007) | .783 (.769-.796) |
| **CHDBioPRS** | | 1.88 (1.75-2.01) | 18.22 | .806 (.007) | .811 (.798-.824) |
| | Q80+ | 2.72 (2.37-3.11) | 14.57 | .791 (.007) | .799 (.786-.812) |
| | Q90+ | 2.64 (2.24-3.09) | 11.79 | .784 (.007) | .792 (.778-.805) |
| | Q95+ | 3.28 (2.70-3.98) | 12.05 | .783 (.007) | .791 (.777-.804) |
| | Q99+ | 4.53 (3.21-6.40) | 8.58 | .776 (.007) | .785 (.771-.798) |

Adjusted for age, gender and PCs1-10

Table S5: FinnGen PRSes CHD survival HR (CI)/1 SD

| PRS | Quant vs 1-Q | HR (CI) | Z | CIndex (SE) | AUC (CI) |
|---|---|---|---|---|---|
| **FinnGen** | | | | | **Baseline .724 (.722-.727)** |
| **BioPRS** | | 1.27 (1.26-1.29) | 39.89 | .696 (.002) | .734 (.732-.737) |
| | Q80+ | 1.50 (1.46-1.55) | 29.29 | .688 (.002) | .730 (.727-.732) |
| | Q90+ | 1.56 (1.51-1.62) | 25.33 | .685 (.002) | .728 (.725-.731) |
| | Q95+ | 1.68 (1.60-1.75) | 22.25 | .683 (.002) | .727 (.724-.730) |
| | Q99+ | 1.84 (1.67-2.02) | 12.60 | .681 (.002) | .725 (.722-.728) |
| **CHDPRS** | | 1.57 (1.55-1.59) | 70.2 | .721 (.002) | .752 (.750-.755) |
| | Q80+ | 2.09 (2.03-2.14) | 54.56 | .703 (.002) | .741 (.738-.743) |
| | Q90+ | 2.23 (2.17-2.31) | 49.69 | .696 (.002) | .737 (.734-.739) |
| | Q95+ | 2.48 (2.38-2.58) | 44.39 | .691 (.002) | .733 (.730-.736) |
| | Q99+ | 3.12 (2.89-3.37) | 28.96 | .683 (.002) | .727 (.724-.730) |
| **CHDBioPRS** | | 1.60 (1.58-1.62) | 73.71 | .725 (.002) | .755 (.752-.758) |
| | Q80+ | 2.21 (2.1752.27) | 59.52 | .706 (.002) | .743 (.740-.746) |
| | Q90+ | 2.32 (2.25-2.40) | 52.42 | .698 (.002) | .738 (.735-.740) |
| | Q95+ | 2.52 (2.43-2.62) | 45.37 | .692 (.002) | .733 (.731-.736) |
| | Q99+ | 3.24 (2.99-3.50) | 29.62 | .684 (.002) | .727 (.724-.730) |

Adjusted for age, gender and PCs1-10

Table S6: Early onset FinnGen PRSes CHD survival HR (CI)/1 SD

| PRS | Quant vs 1-Q | HR (CI) | Z | CIndex (SE) | AUC (CI) |
|---|---|---|---|---|---|
| **FinnGen early onset (<= 55)** | | | | | **Baseline .733 (.727-.739)** |
| **BioPRS** | | 1.51 (1.47-1.55) | 29.04 | .756 (.003) | .759 (.752-.765) |
| | Q80+ | 1.88 (1.77-1.99) | 21.08 | .744 (.003) | .746 (.740-.753) |
| | Q90+ | 2.01 (1.87-2.16) | 19.24 | .741 (.003) | .743 (.737-.750) |
| | Q95+ | 2.20 (2.02-2.41) | 17.18 | .738 (.003) | .741 (.735-.748) |
| | Q99+ | 2.52 (2.11-3.02) | 10.17 | .735 (.003) | .737 (.731-.744) |
| **CHDPRS** | | 2.01 (1.95-2.07) | 47.31 | .781 (.003) | .784 (.778-.791) |
| | Q80+ | 3.05 (2.88-3.23) | 38.25 | .761 (.003) | .764 (.758-.770) |
| | Q90+ | 3.43 (3.22-3.66) | 38.29 | .756 (.003) | .759 (.752-.765) |
| | Q95+ | 3.84 (3.56-4.14) | 35.08 | .749 (.003) | .752 (.745-.758) |
| | Q99+ | 5.34 (4.71-6.06) | 25.98 | .739 (.003) | .742 (.736-.748) |
| **CHDBioPRS** | | 2.10 (2.04-2.16) | 50.58 | .788 (.003) | .791 (.785-.797) |
| | Q80+ | 3.42 (3.24-3.62) | 42.76 | .767 (.003) | .770 (.763-.776) |
| | Q90+ | 3.60 (3.39-3.84) | 40.28 | .757 (.003) | .760 (.753-.766) |
| | Q95+ | 4.04 (3.76-4.35) | 37.08 | .749 (.003) | .752 (.746-.759) |
| | Q99+ | 5.02 (4.40-5.72) | 24.16 | .738 (.003) | .741 (.735-.748) |

Adjusted for age, gender and PCs1-10.
Censored applied for controls at age 55.

Table S7: Early onset UKB Test PRSes CHD survival HR (CI)/1 SD

| PRS | Quant vs 1-Q | HR (CI) | Z | CIndex (SE) | AUC (CI) |
|---|---|---|---|---|---|
| **UKB Test early onset (<= 55)** | | | | | **Baseline .736 (.711-.762)** |
| **BioPRS** | | 1.60 (1.43-1.78) | 8.42 | .763 (.012) | .765 (.740-.788) |
| | Q80+ | 2.34 (1.87-2.94) | 7.34 | .755 (.012) | .757 (.732-.781) |
| | Q90+ | 2.81 (2.17-3.63) | 7.86 | .752 (.013) | .754 (.729-.779) |
| | Q95+ | 2.89 (2.10-3.99) | 6.47 | .746 (.013) | .748 (.723-.772) |
| | Q99+ | 3.31 (1.81-6.04) | 3.90 | .740 (.013) | .741 (.716-.766) |
| **CHDPRS** | | 1.90 (1.69-2.13) | 10.75 | .777 (.012) | .780 (.755-.804) |
| | Q80+ | 2.83 (2.26-3.56) | 8.98 | .766 (.012) | .768 (.744-.792) |
| | Q90+ | 2.91 (2.24-3.78) | 8.00 | .754 (.012) | .756 (.732-.781) |
| | Q95+ | 3.56 (2.62-4.86) | 8.05 | .751 (.012) | .753 (.729-.777) |
| | Q99+ | 5.51 (3.20-9.49) | 6.15 | .741 (.013) | .743 (.718-.768) |
| **CHDBioPRS** | | 2.07 (1.85-2.32) | 12.38 | .788 (.012) | .790 (.766-.814) |
| | Q80+ | 3.51 (2.81-4.39) | 11.03 | .776 (.012) | .778 (.755-.801) |
| | Q90+ | 3.58 (2.79-4.60) | 9.99 | .762 (.012) | .764 (.740-.789) |
| | Q95+ | 4.16 (3.09-5.60) | 9.39 | .755 (.012) | .757 (.732-.782) |
| | Q99+ | 5.15 (3.05-8.70) | 6.13 | .742 (.013) | .743 (.719-.768) |

Adjusted for age, gender and PCs1-10.
Censored applied for controls at age 55.

Table S8: Male UKB Test PRSes CHD survival HR (CI)/1 SD

| PRS | Quant vs 1-Q | HR (CI) | Z | CIndex (SE) | AUC (CI) |
|---|---|---|---|---|---|
| **UKB Test Male** | | | | | **Baseline .710 (.693-.727)** |
| **BioPRS** | | 1.42 (1.32-1.53) | 9.43 | .720 (.009) | .733 (.716-.751) |
| | Q80+ | 1.85 (1.58-2.16) | 7.59 | .711 (.009) | .725 (.707-.742) |
| | Q90+ | 2.12 (1.75-2.56) | 7.76 | .708 (.009) | .722 (.705-.739) |
| | Q95+ | 2.13 (1.66-2.74) | 5.96 | .703 (.009) | .717 (.700-.735) |
| | Q99+ | 2.52 (1.58-4.03) | 3.88 | .698 (.009) | .712 (.695-.730) |
| **CHDPRS** | | 1.72 (1.60-1.86) | 14.47 | .744 (.009) | .757 (.740-.774) |
| | Q80+ | 2.54 (2.18-2.97) | 11.89 | .726 (.009) | .739 (.721-.756) |
| | Q90+ | 2.94 (2.46-3.51) | 11.93 | .721 (.009) | .734 (.717-.751) |
| | Q95+ | 2.79 (2.22-3.52) | 8.74 | .707 (.009) | .721 (.704-.738) |
| | Q99+ | 3.80 (2.53-5.72) | 6.41 | .702 (.009) | .716 (.699-.733) |
| **CHDBioPRS** | | 1.84 (1.70-1.98) | 15.87 | .752 (.009) | .765 (.748-.782) |
| | Q80+ | 2.80 (2.41-3.26) | 13.03 | .733 (.009) | .746 (.729-.764) |
| | Q90+ | 3.03 (2.54-3.61) | 12.36 | .721 (.009) | .734 (.717-.752) |
| | Q95+ | 3.23 (2.59-4.02) | 10.46 | .714 (.009) | .728 (.710-.745) |
| | Q99+ | 4.11 (2.77-6.08) | 7.04 | .703 (.009) | .717 (.699-.734) |

Adjusted for age and PCs1-10.

Table S9: Male FinnGen PRSes CHD survival HR (CI)/1 SD

| PRS | Quant vs 1-Q | HR (CI) | Z | CIndex (SE) | AUC (CI) |
|---|---|---|---|---|---|
| **FinnGen Male** | | | | | **Baseline .571 (.567-.575)** |
| **BioPRS** | | 1.27 (1.25-1.29) | 33.56 | .606 (.002) | .612 (.608-.616) |
| | Q80+ | 1.48 (1.44-1.53) | 24.07 | .593 (.002) | .599 (.595-.603) |
| | Q90+ | 1.56 (1.50-1.63) | 21.62 | .588 (.002) | .596 (.592-.600) |
| | Q95+ | 1.68 (1.59-1.77) | 19.07 | .584 (.002) | .593 (.589-.597) |
| | Q99+ | 1.75 (1.56-1.96) | 9.71 | .578 (.002) | .586 (.582-.590) |
| **CHDPRS** | | 1.58 (1.55-1.60) | 60.06 | .655 (.002) | .652 (.648-.655) |
| | Q80+ | 2.09 (2.03-2.16) | 49.66 | .624 (.002) | .625 (.621-.629) |
| | Q90+ | 2.22 (2.14-2.31) | 41.51 | .608 (.002) | .614 (.610-.618) |
| | Q95+ | 2.45 (2.33-2.57) | 36.57 | .596 (.002) | .603 (.599-.607) |
| | Q99+ | 3.10 (2.83-3.40) | 23.89 | .581 (.002) | .590 (.586-.594) |
| **CHDBioPRS** | | 1.61 (1.59-1.63) | 63.01 | .663 (.002) | .657 (.653-.661) |
| | Q80+ | 2.21 (2.15-2.28) | 50.55 | .629 (.002) | .631 (.627-.635) |
| | Q90+ | 2.33 (2.24-2.41) | 44.33 | .611 (.002) | .616 (.612-.620) |
| | Q95+ | 2.51 (2.40-2.64) | 37.81 | .598 (.002) | .604 (.600-.608) |
| | Q99+ | 2.98 (2.71-3.27) | 22.65 | .581 (.002) | .589 (.585-.593) |

Adjusted for age and PCs1-10.

Table S10: Female FinnGen PRSes CHD survival HR (CI)/1 SD

| PRS | Quant vs 1-Q | HR (CI) | Z | CIndex (SE) | AUC (CI) |
|---|---|---|---|---|---|
| **FinnGen Female** | | | | | **Baseline .710 (.705-.710)** |
| **BioPRS** | | 1.27 (1.24-1.30) | 20.97 | .625 (.004) | .720 (.715-.725) |
| | Q80+ | 1.50 (1.42-1.57) | 15.43 | .611 (.004) | .715 (.710-.721) |
| | Q90+ | 1.59 (1.49-1.69) | 13.95 | .607 (.004) | .714 (.709-.719) |
| | Q95+ | 1.67 (1.53-1.82) | 11.68 | .604 (.004) | .713 (.708-.718) |
| | Q99+ | 1.88 (1.57-2.24) | 6.98 | .598 (.004) | .711 (.706-.716) |
| **CHDPRS** | | 1.53 (1.50-1.57) | 35.65 | .657 (.004) | .738 (.733-.743) |
| | Q80+ | 2.05 (1.95-2.15) | 30.04 | .638 (.004) | .728 (.723-.733) |
| | Q90+ | 2.24 (2.12-2.38) | 27.00 | .629 (.004) | .725 (.720-.730) |
| | Q95+ | 2.51 (2.34-2.70) | 24.84 | .619 (.004) | .722 (.717-.727) |
| | Q99+ | 3.20 (2.80-3.66) | 17.06 | .604 (.004) | .715 (.710-.720) |
| **CHDBioPRS** | | 1.56 (1.53-1.60) | 37.46 | .665 (.005) | .741 (.736-.746) |
| | Q80+ | 2.14 (2.03-2.24) | 30.35 | .645 (.004) | .730 (.725-.735) |
| | Q90+ | 2.29 (2.16-2.42) | 27.81 | .631 (.004) | .726 (.721-.731) |
| | Q95+ | 2.50 (2.32-2.69) | 24.52 | .621 (.004) | .722 (.717-.727) |
| | Q99+ | 3.70 (3.23-4.23) | 19.08 | .605 (.004) | .715 (.710-.720) |

Adjusted for age and PCs1-10.

Table S11: Female UKB Test PRSes CHD survival HR (CI)/1 SD

| PRS | Quant vs 1-Q | HR (CI) | Z | CIndex (SE) | AUC (CI) |
|---|---|---|---|---|---|
| **UKB Test Female** | | | | | **Baseline .726 (.694-.757)** |
| **BioPRS** | | 1.53 (1.34-1.75) | 6.14 | .746 (.015) | .750 (.719-.780) |
| | Q80+ | 1.75 (1.31-2.35) | 3.75 | .731 (.016) | .736 (.705-.767) |
| | Q90+ | 2.11 (1.48-2.99) | 4.16 | .731 (.016) | .735 (.703-.766) |
| | Q95+ | 2.43 (1.56-3.78) | 3.93 | .730 (.016) | .734 (.703-.765) |
| | Q99+ | 2.72 (1.12-6.63) | 2.21 | .722 (.016) | .727 (.695-.758) |
| **CHDPRS** | | 1.72 (1.50-1.98) | 7.59 | .756 (.015) | .760 (.730-.790) |
| | Q80+ | 2.16 (1.62-2.88) | 5.24 | .735 (.016) | .739 (.708-.770) |
| | Q90+ | 2.40 (1.71-3.37) | 5.03 | .731 (.016) | .736 (.704-.767) |
| | Q95+ | 3.86 (2.66-5.59) | 7.12 | .737 (.016) | .741 (.709-.772) |
| | Q99+ | 3.81 (1.86-7.80) | 3.65 | .728 (.016) | .732 (.701-.763) |
| **CHDBioPRS** | | 1.86 (1.62-2.13) | 8.78 | .766 (.015) | .770 (.741-.799) |
| | Q80+ | 2.78 (2.11-3.66) | 7.22 | .756 (.015) | .756 (.726-.786) |
| | Q90+ | 2.72 (1.96-3.79) | 5.93 | .736 (.016) | .741 (.709-.772) |
| | Q95+ | 4.03 (2.77-5.85) | 7.32 | .739 (.016) | .743 (.712-.775) |
| | Q99+ | 5.15 (2.77-9.58) | 5.18 | .726 (.016) | .730 (.698-.761) |

Adjusted for age and PCs1-10.

Table S12: Female Early onset FinnGen PRSes CHD survival HR (CI)/1 SD

| **Finngen Female early onset <= 60** | | | | | **Baseline .636 (.625-.646)** |
|---|---|---|---|---|---|
| **BioPRS** | | 1.44 (1.38-1.51) | 15.69 | .668 (.006) | .670 (.659-.681) |
| | Q80+ | 1.82 (1.65-2.00) | 12.00 | .655 (.006) | .657 (.651-.668) |
| | Q90+ | 1.93 (1.72-2.17) | 10.89 | .650 (.006) | .651 (.640-.663) |
| | Q95+ | 2.03 (1.74-2.36) | 9.08 | .645 (.006) | .646 (.635-.658) |
| | Q99+ | 2.08 (1.51-2.86) | 4.52 | .639 (.006) | .640 (.629-.652) |
| **CHDPRS** | | 1.82 (1.74-1.90) | 24.68 | .697 (.006) | .698 (.686-.710) |
| | Q80+ | 2.64 (2.41-2.91) | 20.16 | .676 (.006) | .677 (.666-.689) |
| | Q90+ | 2.90 (2.61-3.23) | 19.68 | .668 (.006) | .670 (.658-.681) |
| | Q95+ | 3.40 (3.00-3.85) | 19.15 | .659 (.006) | .661 (.649-.672) |
| | Q99+ | 4.90 (3.99-6.02) | 15.16 | .647 (.006) | .649 (.637-.660) |
| **CHDBioPRS** | | 1.90 (1.81-1.99) | 26.51 | .707 (.006) | .709 (.697-.720) |
| | Q80+ | 2.97 (2.71-3.26) | 22.92 | .684 (.006) | .686 (.674-.698) |
| | Q90+ | 3.06 (2.75-3.39) | 20.93 | .671 (.006) | .673 (.661-.684) |
| | Q95+ | 3.39 (3.00-3.84) | 19.17 | .662 (.006) | .663 (.652-.675) |
| | Q99+ | 4.80 (3.88-5.92) | 14.56 | .646 (.006) | .647 (.636-.658) |

Adjusted for age and PCs1-10.
Censored applied for controls at age 60.

Table S13: Male Early onset FinnGen PRSes CHD survival HR (CI)/1 SD

| PRS | Quant vs 1-Q | HR (CI) | Z | CIndex (SE) | AUC (CI) |
|---|---|---|---|---|---|
| **Finngen Male early onset <= 50** | | | | | **Baseline .578 (.567-.590)** |
| **BioPRS** | | 1.53 (1.47-1.60) | 18.95 | .639 (.006) | .641 (.629-.653) |
| | Q80+ | 2.02 (1.85-2.22) | 15.00 | .611 (.006) | .613 (.600-.625) |
| | Q90+ | 2.09 (1.87-2.33) | 13.01 | .597 (.006) | .598 (.586-.611) |
| | Q95+ | 2.31 (2.01-2.66) | 11.84 | .593 (.006) | .594 (.582-.607) |
| | Q99+ | 2.44 (1.85-3.24) | 6.23 | .584 (.006) | .585 (.572-.597) |
| **CHDPRS** | | 2.17 (2.07-2.27) | 32.95 | .706 (.006) | .708 (.697-.720) |
| | Q80+ | 3.36 (3.07-3.68) | 26.03 | .657 (.006) | .658 (.646-.671) |
| | Q90+ | 3.98 (3.61-4.39) | 27.09 | .642 (.006) | .644 (.631-.656) |
| | Q95+ | 4.66 (4.16-5.20) | 27.02 | .624 (.006) | .626 (.613-.638) |
| | Q99+ | 5.88 (4.88-7.09) | 18.58 | .593 (.006) | .594 (.582-.607) |
| **CHDBioPRS** | | 2.26 (2.16-2.36) | 34.74 | .716 (.006) | .719 (.707-.730) |
| | Q80+ | 3.80 (3.47-4.15) | 29.17 | .670 (.006) | .672 (.659-.684) |
| | Q90+ | 4.07 (3.70-4.48) | 32.47 | .642 (.006) | .644 (.631-.657) |
| | Q95+ | 4.76 (4.26-5.32) | 27.07 | .625 (.006) | .626 (.614-.639) |
| | Q99+ | 5.63 (4.66-6.82) | 20.40 | .591 (.006) | .592 (.580-.605) |

Adjusted for age and PCs1-10.
Censored applied for controls at age 50.

Table S14: Female Early onset UKB Test PRSes CHD survival HR (CI)/1 SD

| PRS | Quant vs 1-Q | HR (CI) | Z | CIndex (SE) | AUC (CI) |
|---|---|---|---|---|---|
| **UKB Test Female early onset <= 60** | | | | | **Baseline .654 (.609-.699)** |
| **BioPRS** | | 1.57 (1.31-1.89) | 4.89 | .701 (.022) | .702 (.659-.746) |
| | Q80+ | 1.90 (1.29-2.79) | 3.26 | .676 (.023) | .677 (.631-.723) |
| | Q90+ | 2.19 (1.39-3.45) | 3.36 | .668 (.023) | .669 (.622-.715) |
| | Q95+ | 2.36 (1.33-4.21) | 2.92 | .662 (.023) | .663 (.619-.707) |
| | Q99+ | 1.62 (0.40-6.57) | 0.67 | .655 (.023) | .656 (.611-.702) |
| **CHDPRS** | | 1.72 (1.52-2.15) | 5.61 | .715 (.021) | .717 (.674-.759) |
| | Q80+ | 2.07 (1.41-3.04) | 3.71 | .681 (.023) | .682 (.636-.728) |
| | Q90+ | 2.48 (1.59-3.86) | 4.00 | .677 (.024) | .678 (.631-.724) |
| | Q95+ | 3.81 (2.33-6.25) | 5.31 | .681 (.023) | .682 (.636-.728) |
| | Q99+ | 4.30 (1.72-10.74) | 3.12 | .666 (.022) | .667 (.624-.711) |
| **CHDBioPRS** | | 1.89 (1.57-2.27) | 6.66 | .732 (.021) | .733 (.691-.775) |
| | Q80+ | 3.18 (2.21-4.59) | 6.20 | .715 (.023) | .715 (.671-.760) |
| | Q90+ | 2.65 (1.71-4.10) | 4.37 | .681 (.023) | .681 (.635-.728) |
| | Q95+ | 3.92 (2.40-6.40) | 5.46 | .683 (.023) | .684 (.638-.730) |
| | Q99+ | 4.03 (1.61-10.13) | 2.97 | .663 (.023) | .664 (.619-.709) |

Adjusted for age and PCs1-10.
Censored applied for controls at age 60.

Table S15: Male Early onset UKB Test PRSes CHD survival HR (CI)/1 SD

| PRS | Quant vs 1-Q | HR (CI) | Z | CIndex (SE) | AUC (CI) |
|---|---|---|---|---|---|
| **UKB Test Male early onset <= 50** | | | | | **Baseline .612 (.563-.660)** |
| **BioPRS** | | 1.36 (1.15-1.62) | 3.50 | .636 (.025) | .637 (.588-.686) |
| | Q80+ | 1.66 (1.13-2.44) | 2.60 | .629 (.024) | .630 (.582-.677) |
| | Q90+ | 2.70 (1.78-4.09) | 4.68 | .645 (.025) | .645 (.596-.695) |
| | Q95+ | 2.53 (1.48-4.35) | 3.38 | .625 (.025) | .626 (.577-.675) |
| | Q99+ | 4.82 (2.12-10.94) | 3.75 | .621 (.025) | .622 (.572-.672) |
| **CHDPRS** | | 1.81 (1.52-2.15) | 6.62 | .675 (.025) | .676 (.627-.725) |
| | Q80+ | 2.75 (1.91-3.92) | 5.49 | .665 (.024) | .667 (.618-.715) |
| | Q90+ | 3.40 (2.28-5.05) | 6.04 | .657 (.025) | .657 (.606-.707) |
| | Q95+ | 4.04 (2.54-6.42) | 5.91 | .637 (.026) | .638 (.587-.689) |
| | Q99+ | 9.00 (4.70-17.21) | 6.64 | .629 (.025) | .630 (.579-.680) |
| **CHDBioPRS** | | 1.87 (1.56-2.23) | 6.86 | .687 (.025) | .688 (.639-.737) |
| | Q80+ | 2.73 (1.91-3.92) | 5.49 | .670 (.024) | .671 (.624-.718) |
| | Q90+ | 3.66 (2.48-5.41) | 6.53 | .673 (.025) | .674 (.625-.723) |
| | Q95+ | 5.10 (3.30-7.88) | 7.33 | .664 (.026) | .664 (.614-.715) |
| | Q99+ | 7.94 (4.02-15.68) | 5.97 | .633 (.006) | .634 (.583-.684) |

Adjusted for age and PCs1-10.
Censored applied for controls at age 50.
