## Supplemental Figures 1-3 for "Integration of biomarker polygenic risk score improves prediction of coronary heart disease in UK Biobank and FinnGen"

A. Glmnet (elastic net regression) with an alpha value of 0.50 found 10 biomarkers in its optimal model (lambda.1SE setting), as indicated by the plot's right vertical dashed line and the selected inversed normalize (*_int) biomarker predictors are shown on the right. These same 10 biomarkers were found to be the optimal model using alpha value of 0.25. A lambda.min setting for both 0.25 and 0.50 lambda values selected all 16 biomarkers (as indicated by the left vertical dashed line) is not selected due to overfitting.

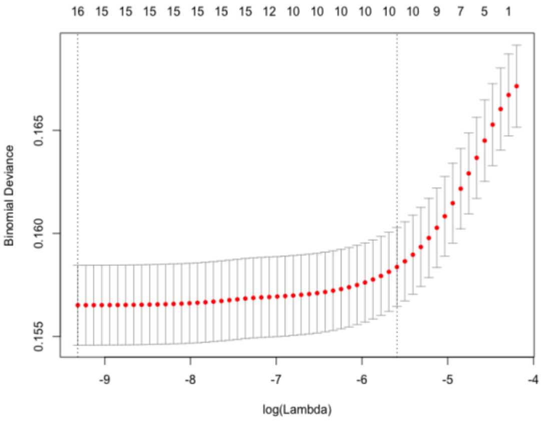


| | |
|---|---|
| adj_hdl_int | -0.16225960 |
| adj_ldl_int | 0.11438159 |
| adj_tg_int | 0.04423273 |
| adj_tc1_int | . |
| bmi_int | . |
| ApoB_int | 0.08207446 |
| ApoA1_int | -0.02131672 |
| gluc_int | . |
| lpa_int | . |
| t2d_int | . |
| sbp_int | 0.25952935 |
| dbp_int | . |
| crea_int | 0.20981747 |
| CRP_int | 0.03610340 |
| hba1c_int | 0.06288727 |
| cigs_perday_int | 0.08135247 |

B. Glmnet with an alpha value of 0.75 found 9 biomarkers (ApoA1 excluded). To balance sensitivity and fit, we select the 10 biomarker optimal models, selected by models with 0.50 and 0.25 alpha values.

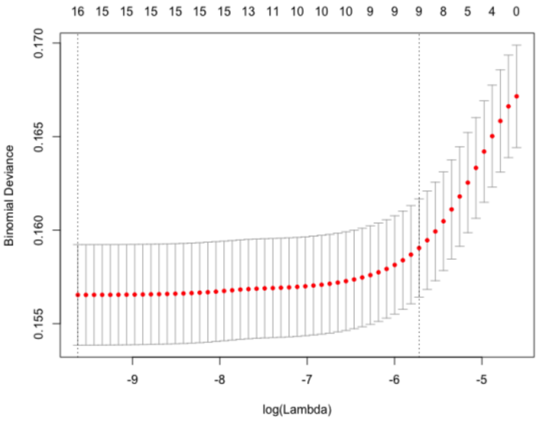


| | |
|---|---|
| adj_hdl_int | -0.17227396 |
| adj_ldl_int | 0.09088489 |
| adj_tg_int | 0.03217825 |
| adj_tc1_int | . |
| bmi_int | . |
| ApoB_int | 0.08156334 |
| ApoA1_int | . |
| gluc_int | . |
| lpa_int | . |
| t2d_int | . |
| sbp_int | 0.24881371 |
| dbp_int | . |
| crea_int | 0.18067260 |
| CRP_int | 0.01205109 |
| hba1c_int | 0.04223650 |
| cigs_perday_int | 0.06876118 |

**Figure S1 CHD biomarkers**. Optimal CHD biomarkers are selected with Glmnet (elastic net) regression model optimal (lambda.1se) mode setting.

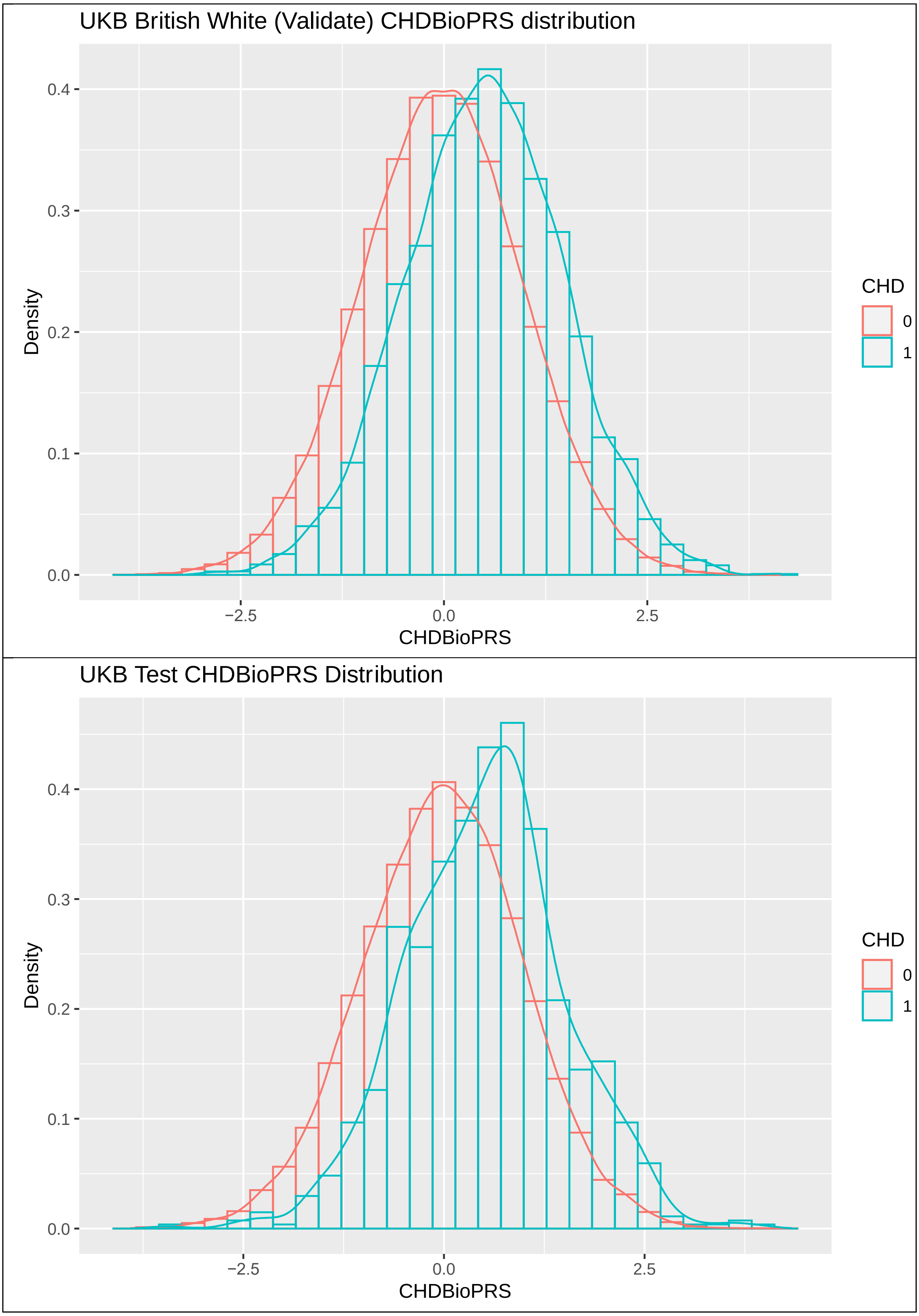


**Figure S2 CHDBioPRS score distribution in cases and controls.** CHDBioPRS scores for UKB Training (not shown), Validation (top) and Test (bottom) cohorts, are approximately normally distributed and on average, higher in CHD cases (blue) than in controls (red).

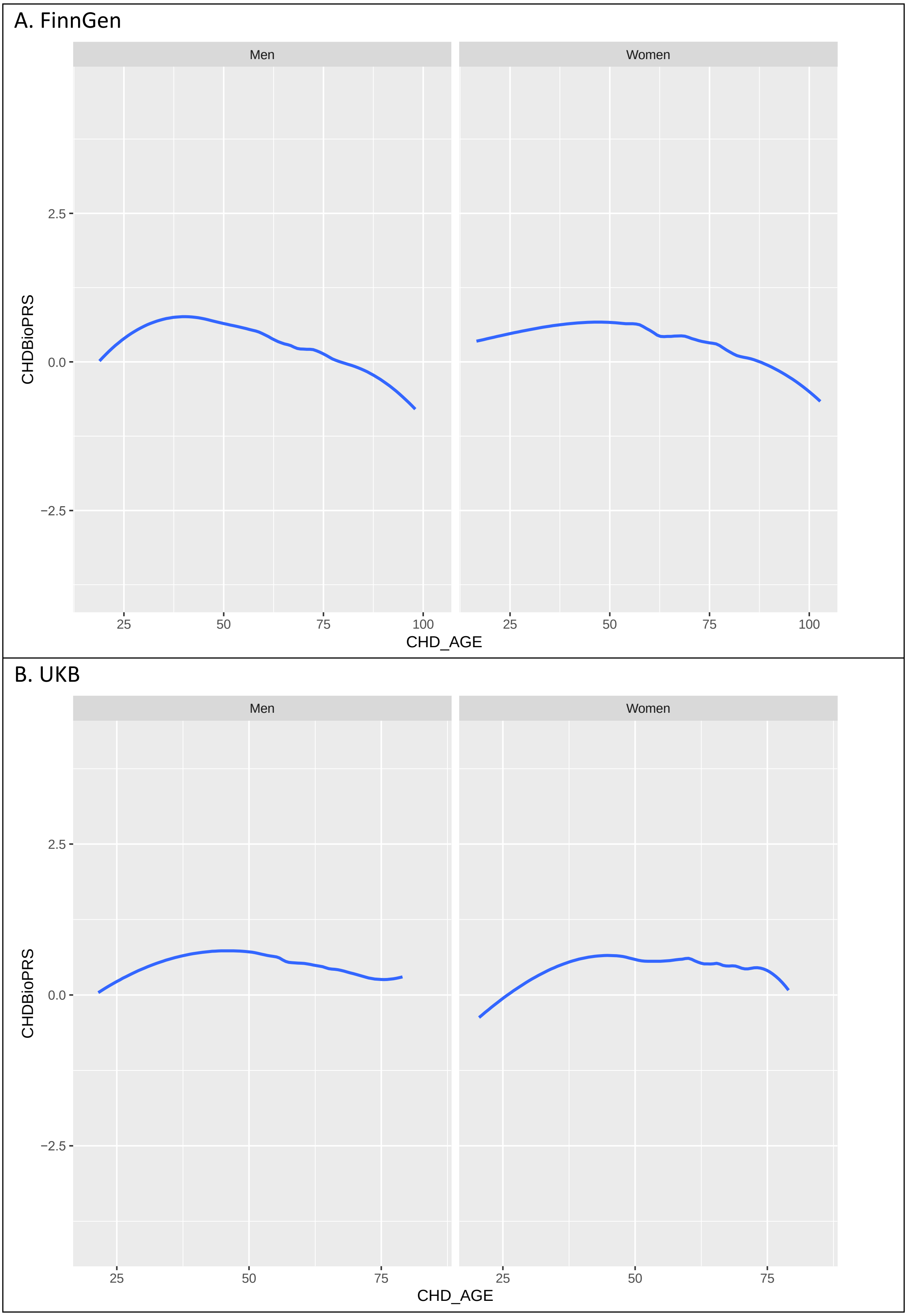


**Figure S3. LOESS regression curves of the mean CHDBioPRS scores and onset age separated by sex and cohorts (A) FinnGen and (B) UKB.**
